# Quantification of Fundus Autofluorescence Features in a Molecularly Characterized Cohort of More Than 3500 Inherited Retinal Disease Patients from the United Kingdom

**DOI:** 10.1101/2024.03.24.24304809

**Authors:** William Woof, Thales A. C. de Guimarães, Saoud Al-Khuzaei, Malena Daich Varela, Sagnik Sen, Pallavi Bagga, Bernardo Mendes, Mital Shah, Paula Burke, David Parry, Siying Lin, Gunjan Naik, Biraja Ghoshal, Bart Liefers, Dun Jack Fu, Michalis Georgiou, Quang Nguyen, Alan Sousa da Silva, Yichen Liu, Yu Fujinami-Yokokawa, Dayyanah Sumodhee, Praveen Patel, Jennifer Furman, Ismail Moghul, Mariya Moosajee, Juliana Sallum, Samantha R. De Silva, Birgit Lorenz, Frank Holz, Kaoru Fujinami, Andrew R Webster, Omar Mahroo, Susan M. Downes, Savita Madhusudhan, Konstantinos Balaskas, Michel Michaelides, Nikolas Pontikos

**Author notes:** Authors contributing equally (William Woof, Thales A. C. de Guimarães, Saoud Al-Khuzaei, Malena Daich Varela, Sagnik Sen, Konstantinos Balaskas, Michel Michaelides, Nikolas Pontikos).

## Abstract

**Purpose:** To quantify relevant fundus autofluorescence (FAF) image features cross-sectionally and longitudinally in a large cohort of inherited retinal diseases (IRDs) patients.

**Design:** Retrospective study of imaging data (55-degree blue-FAF on Heidelberg Spectralis) from patients.

**Participants:** Patients with a clinical and molecularly confirmed diagnosis of IRD who have undergone FAF 55-degree imaging at Moorfields Eye Hospital (MEH) and the Royal Liverpool Hospital (RLH) between 2004 and 2019.

**Methods:** Five FAF features of interest were defined: vessels, optic disc, perimacular ring of increased signal (ring), relative hypo-autofluorescence (hypo-AF) and hyper-autofluorescence (hyper-AF). Features were manually annotated by six graders in a subset of patients based on a defined grading protocol to produce segmentation masks to train an AI model, AIRDetect, which was then applied to the entire MEH imaging dataset.

**Main Outcome Measures:** Quantitative FAF imaging features including area in mm^2^ and vessel metrics, were analysed cross-sectionally by gene and age, and longitudinally to determine rate of progression. AIRDetect feature segmentation and detection were validated with Dice score and precision/recall, respectively.

**Results:** A total of 45,749 FAF images from 3,606 IRD patients from MEH covering 170 genes were automatically segmented using AIRDetect. Model-grader Dice scores for disc, hypo-AF, hyper-AF, ring and vessels were respectively 0.86, 0.72, 0.69, 0.68 and 0.65. The five genes with the largest hypo-AF areas were *CHM*, *ABCC6*, *ABCA4*, *RDH12*, and *RPE65*, with mean per-patient areas of 41.5, 30.0, 21.9, 21.4, and 15.1 mm^2^. The five genes with the largest hyper-AF areas were *BEST1*, *CDH23*, *RDH12*, *MYO7A*, and *NR2E3*, with mean areas of 0.49, 0.45, 0.44, 0.39, and 0.34 mm^2^ respectively. The five genes with largest ring areas were *CDH23*, *NR2E3*, *CRX*, *EYS* and *MYO7A,* with mean areas of 3.63, 3.32, 2.84, 2.39, and 2.16 mm^2^. Vessel density was found to be highest in *EFEMP1*, *BEST1*, *TIMP3*, *RS1*, and *PRPH2* (10.6%, 10.3%, 9.8%, 9.7%, 8.9%) and was lower in Retinitis Pigmentosa (RP) and Leber Congenital Amaurosis genes. Longitudinal analysis of decreasing ring area in four RP genes (*RPGR, USH2A, RHO, EYS*) found *EYS* to be the fastest progressor at -0.18 mm^2^/year.

**Conclusions:** We have conducted the first large-scale cross-sectional and longitudinal quantitative analysis of FAF features across a diverse range of IRDs using a novel AI approach.

## Introduction

Inherited retinal diseases (IRDs) are clinically and genetically heterogeneous disorders that affect the retina and represent the leading cause of legal blindness among working-age adults in England and Wales, and the second commonest cause in childhood ^1^. A recent study from Austria, covering nine federal states, has also shown that IRDs are now the leading cause of registered blindness in Austrian children and working-age adults^2^. This group of disorders can be caused by genetic variants in any one of over 300 genes ^3–5^.

Many IRDs are associated with structural changes within the retina, which can be detected with retinal imaging using different imaging modalities such as colour fundus, infrared-reflectance (IR), spectral-domain optical coherence tomography (SD-OCT), or fundus autofluorescence (FAF). FAF is of particular importance in the context of IRDs, as it allows the detection of patterns of fluorophores, often at the level of the photoreceptors and retinal pigment epithelium (RPE), which can be indicative of pathological changes such as loss of overlying photoreceptors ^6,7^. Some of these FAF signal changes are highly characteristic of specific IRDs and can indicate features such as areas of RPE atrophy or lipofuscin deposits. FAF is listed as a primary or secondary outcome in multiple clinical trials, and it has become a useful retinal biomarker for diagnostic and prognostication purposes in a wide variety of IRDs ^4,6,8,9^.

The identification and quantification of disease-associated features within retinal imaging is critical for diagnosis, monitoring disease progression, providing prognostic information and assessing treatments in IRDs. The first steps in quantifying retinal imaging-based biomarkers of disease involves identification and segmentation of these features. Manual segmentation performed by human annotators is time-consuming and requires expert annotators, which makes this process subjective and not feasible on a large scale. Automated identification and segmentation of IRD features in a reliable way is important for enabling the routine use of these data quantitatively in clinical practice and to help further our understanding of these diseases.

Existing studies that have used deep learning to segment IRD features from retinal images have so far focused on specific IRD phenotypes such as retinitis pigmentosa (RP), Stargardt (STGD1), and choroideremia (CHM) ^10,11^.

To support our analysis on a broad range of different IRD phenotypes, we developed AIRDetect, a deep learning model that can automatically identify and segment relevant features from FAF images. We apply AIRDetect to the entire cohort of IRD patients with molecularly confirmed diagnoses at Moorfields Eye Hospital (MEH), to identify genotype-phenotype associations, as well as quantify disease progression.

## Methods

### Dataset Curation

Patients’ genotypes were extracted from the Genetics database of MEH (London, UK) ^3,12^. Patients’ images were exported from the Heidelberg Imaging (Heyex) database (Heidelberg Engineering, Heidelberg, Germany) based on their hospital number, for records between 2004-06-17 and 2019-10-22. All 55-degree FAF images were 488nm blue-FAF images captured by the Heidelberg Spectralis and the HRA2 imaging platforms.

A dataset of 736 blue-FAF images (55-degree) from 573 patients from MEH were annotated with four different image features, optic disc, regions of hyper- and hypo-autofluorescence (AF), and perimacular ring of increased signal, and a further set of 206 blue-FAF images (55-degree) from 127 patients from the Royal Liverpool Hospital (RLH) were annotated with the retina vessel tree. A grading protocol was defined for IRD retinal feature annotations (**Table 1)** ^13^. The Dice similarity coefficient score was used to assess inter-grader agreement^14^. The Dice similarity coefficient is defined as twice the area of overlap between two annotations divided by the total area occupied by the two annotations. It ranges from one for perfect overlap between two annotations to zero for no overlap between two annotations. The intergrader agreement was not found to be significantly different between the graders. Manual grading was completed over an 18-month period from June 2022 to December 2023 by four graders, with two additional graders carrying out the vessel segmentation at RLH. The four MEH graders were research fellows with over 5 years’ experience in medical retina, three of which had 3 years’ experience with FAF scans and IRDs. The two RLH graders were staff from the RLH Reading Centre with over 5 years’ experience in vessel annotation on FAF scans. Manual grading was performed using the Moorfields Grading Portal online platform (grading.readingcentre.org). A full breakdown of the manually annotated dataset is given in **Supplementary Table 1**.

**Table 1:**
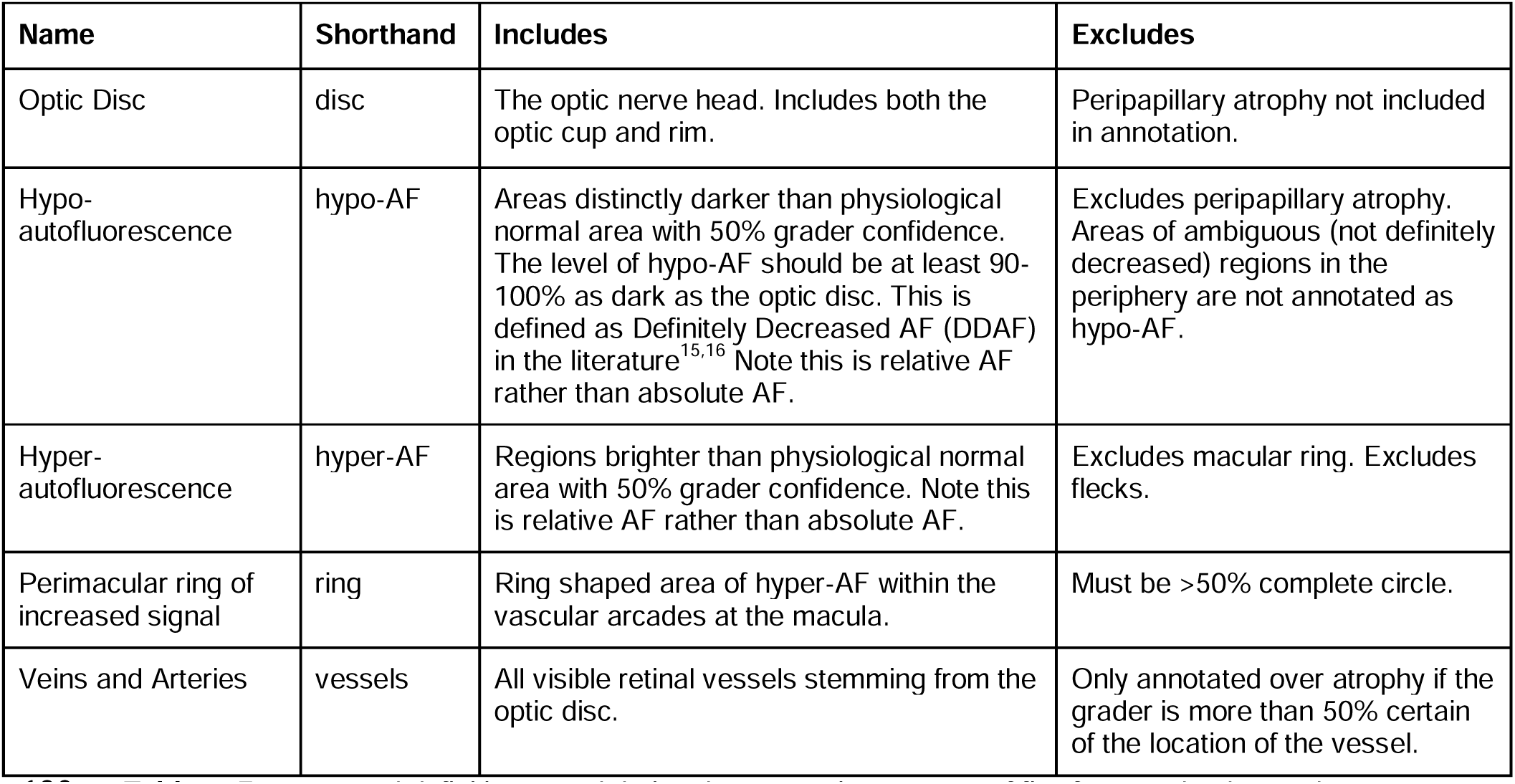
Features and definitions used during the annotation process of five features by the graders.

### Training and Test Datasets

The annotated dataset was compiled, and any images without confirmation for all features from at least one grader at the time of model development were discarded, and, to avoid bias, the annotation from a single grader was randomly selected where multiple grader annotations were available for a single image. After this process there were 554 images from 464 patients from MEH. The MEH training set consisted of 506 images from 424 patients. The MEH hold-out test set consisted of 48 images from 40 patients. The RLH training set consisted of 72 images from 52 patients from RLH. The RLH hold-out test set consisted of 23 images from 22 patients. Training sets were split into five separate sets for use with 5-fold cross validation, ensuring a balanced representation of each class across folds. Assignment to the training and test sets was done at patient-level to avoid any potential data leakage. The data flowchart is fully described in **Supplementary Figure 1**.

### Development of AIRDetect Segmentation Model

For training the AIRDetect segmentation model, we selected the nnU-Net (no-new-UNet) framework for its adaptability and performance in automatic medical image segmentation tasks ^17^. At its core, nnU-Net leverages a fully convolutional network design inspired by the U-Net architecture, renowned for its efficacy in medical imaging tasks ^18–20^. The overlying nnU-Net framework then automatically configures its network architecture, preprocessing, and training strategy based on the dataset’s characteristics, optimising for performance, without requiring manual hyperparameter tuning or architecture modifications from the user.

For the five different image features, we trained two separate nnU-net models. A single multi-class model for disc, hyper-AF and hypo-AF, and ring, and a separate single-class model for vessels. As with common practice for nn-Unet each model consisted of an ensemble of five U-nets with identical architectures, but different weights, trained independently and then ensembled at inference, taking the unweighted average of the probability scores across networks.

The model was trained using a sum of Dice and cross-entropy loss functions to optimise for multi-class segmentation accuracy. Hyperparameters, such as learning rate and batch size, were selected by the nnU-Net based on its analysis of the dataset. Training was curtailed at 200 epochs as this was sufficient to achieve convergence in most cases.

### Validation of AIRDetect Segmentation Model

Model validation was assessed using the Dice coefficient between the model predictions and the corresponding grader annotation on the hold-out test set. Where images were double-graded, we took the mean of the model-grader Dice for each grading. We also analysed the accuracy of the model-grader agreement for simple presence/absence detection where we counted cases as positive for which the model/annotator marked at least some part of the image for the given feature, and negative otherwise, from which we derived presence/absence detection accuracy, precision and recall.

### Automatic Annotations on Real World IRD Dataset

The trained models were applied to automatically segment 45,749 FAF images (55-degree) from 3,606 IRD patients with a molecularly confirmed diagnosis from MEH covering 170 genes ^3,12^. This took on average 1 second per image parallelised over four 3090 Nvidia GPUs amounting to approximately 3-4 hours in total. In comparison a human grader could take 5-30 minutes per scans amounting to 5-3 months in total. Images where the optic disc was not segmented by the model were removed, as these images were of poor quality or not centred on the macula (**Supplementary Figure 2**). Results were analysed from 33,042 FAF images from 3,496 patients, after filtering.

For each of the generated masks we extracted: a) if the feature was present or absent; b) the area, number of pixels in the segmented mask multiplied by the resolution; c) the number of connected components, found using watershed clustering ^21^; d) feature brightness, mean intensity of pixels from the region covered by the segmented mask. For vessels, we calculated a selection of metrics defined in **Supplementary Table 2**, using the provided code from the reti-py library as used in the AutoMorph repository ^22^. Features were also analysed based on their distance from the fovea.

To calculate rate of progression for a given feature, a linear regression was fit to each patient-eye, taking time since the first appointment (in years) as the independent variable, and taking the calculated areas of the segmented feature at each time-point as the measured variable. The slope of the regression was then averaged across eyes per-patient to give a rate of progression. Where multiple scans per eye were present for a given date, we took the most recent scan with the rationale that good quality scans were less likely to lead to further imaging by the operator.

## Results

### AIRDetect Model Validation

Examples of AIRDetect segmentation output are presented in **Figure 1**. Model-grader Dice scores for disc, hypo-AF, hyper-AF, ring and vessels were respectively 0.86, 0.72, 0.69, 0.68 and 0.65, with intergrader Dice scores of 0.82, 0.75, 0.72, 0.80, 0.95, respectively. Model detection accuracy ranged from 77% to 83% (excluding anatomical features) (**Table 2)**. Features which were the most challenging to detect were hyper-AF and ring as those had the lowest precision scores at 0.53 and 0.60 respectively.

**Figure 1:**
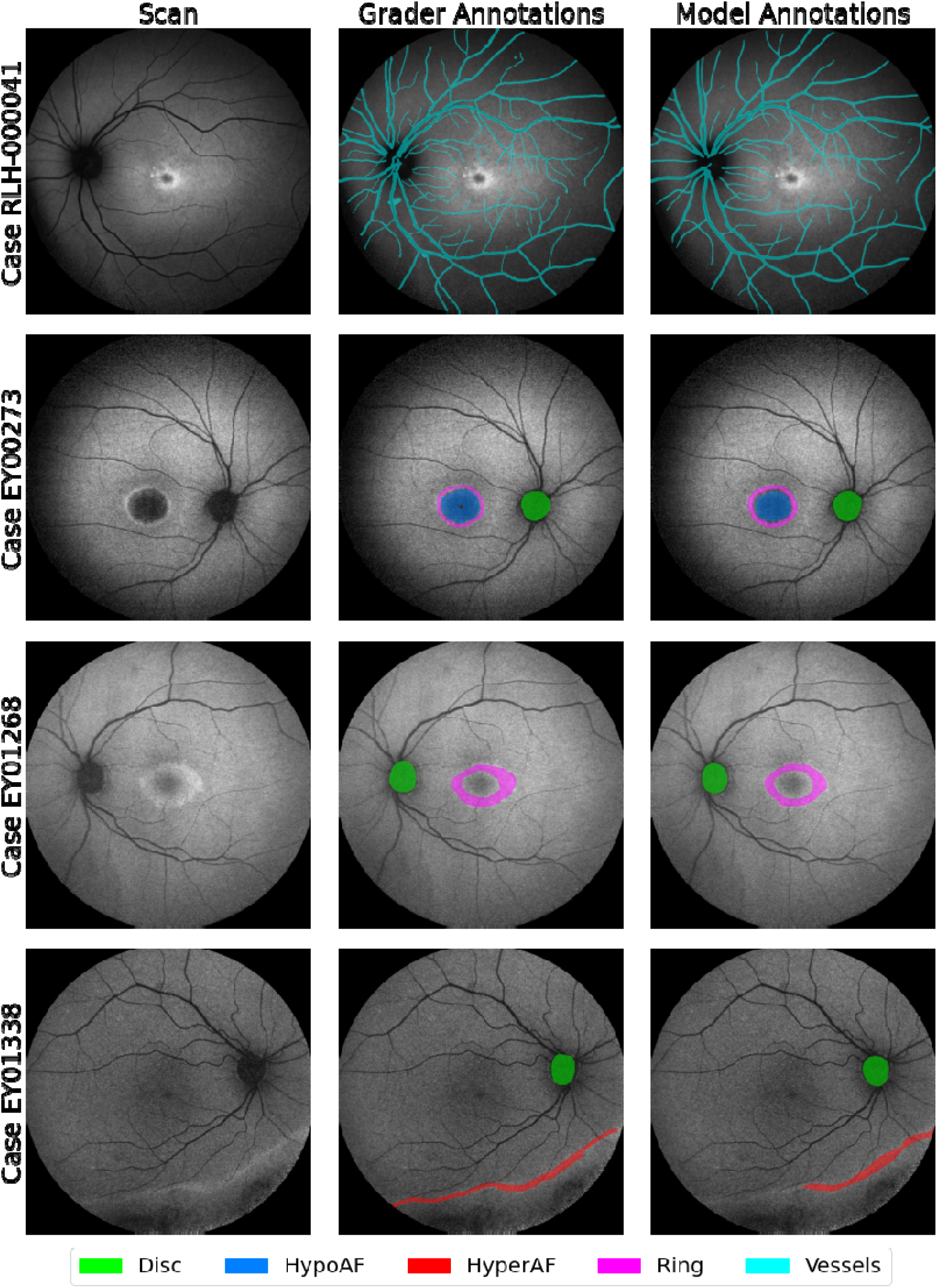
Examples of manually and automatically segmented masks for the five features: vessels, disc, ring, hyper- and hypo-autofluorescence. The vessel dataset was separate to the rest of the data, so vessel visualisation is separate from other features.

**Table 2:** Segmentation model training data and results. Dice score quantifies the model’s segmentation performance and presence/absence quantifies its feature detection performance. Total = number of annotated images. Incidence = percent of images with gradable feature. Dice inter-grader = inter-grader agreement of double-graded images (repeated from **Table 2** for reference). Dice model-grader = Dice score between model and graders, with mean scores used when images were double-graded.

| Feature | Train set |  | Test Set |  | Segmentation (Dice) |  | Detection (Presence/Absence) |  |  |
| --- | --- | --- | --- | --- | --- | --- | --- | --- | --- |
|  | Total | Incidence | Total | Incidence | Inter-grader | Model-grader | Accuracy | Precision | Recall |
| disc | 506 | 98% | 48 | 98% | 0.82 | 0.86 | - | - | - |
| hypo-AF | 506 | 70% | 48 | 44% | 0.75 | 0.72 | 83.3% | 0.81 | 0.81 |
| hyper-AF | 506 | 18% | 48 | 23% | 0.72 | 0.69 | 79.2% | 0.53 | 0.82 |
| ring | 506 | 32% | 48 | 31% | 0.80 | 0.68 | 77.1% | 0.60 | 0.80 |
| vessels | 72 | 100% | 23 | 100% | 0.94 | 0.65 | - | - | - |

### Genotype-phenotype Associations

Analysing associations between identified features and genes across most common genes (**Supplementary Table 3**), the five genes with the largest hypo-AF areas were *CHM*, *ABCC6*, *ABCA4*, *RDH12*, and *RPE65*, with mean per-patient areas of 41.5, 30.0, 21.9, 21.4, and 15.1 mm^2^ (**Figure 2a)**. The five genes with the largest hyper-AF areas were *BEST1*, *CDH23*, *RDH12*, *MYO7A*, and *NR2E3*, with mean areas of 0.49, 0.45, 0.44, 0.39, and 0.34 mm^2^ respectively (**Figure 2b**). The five genes with largest ring areas were *CDH23*, *NR2E3*, *CRX*, *EYS* and *MYO7A,* with mean areas of 3.63, 3.32, 2.84, 2.39, and 2.16 mm^2^ (**Figure 2c**). At the gene variant level, *ABCA4* p.(Gly1961Glu) showed a higher ring area than other common *ABCA4* variants (**Supplementary Figure 4).** Vessel density was found to be highest in *EFEMP1*, *BEST1*, *TIMP3*, *RS1*, and *PRPH2* (10.6%, 10.3%, 9.8%, 9.7%, 8.9%) and was lower in Retinitis Pigmentosa (RP) and Leber Congenital Amaurosis associated genes (**Figure 2d).** A full breakdown of features across the 30 most common genes is given in **Supplementary Table 3**, for all genes in **Supplementary Table 4** and for vessels in **Supplementary Table 5**.

**Figure 2:**
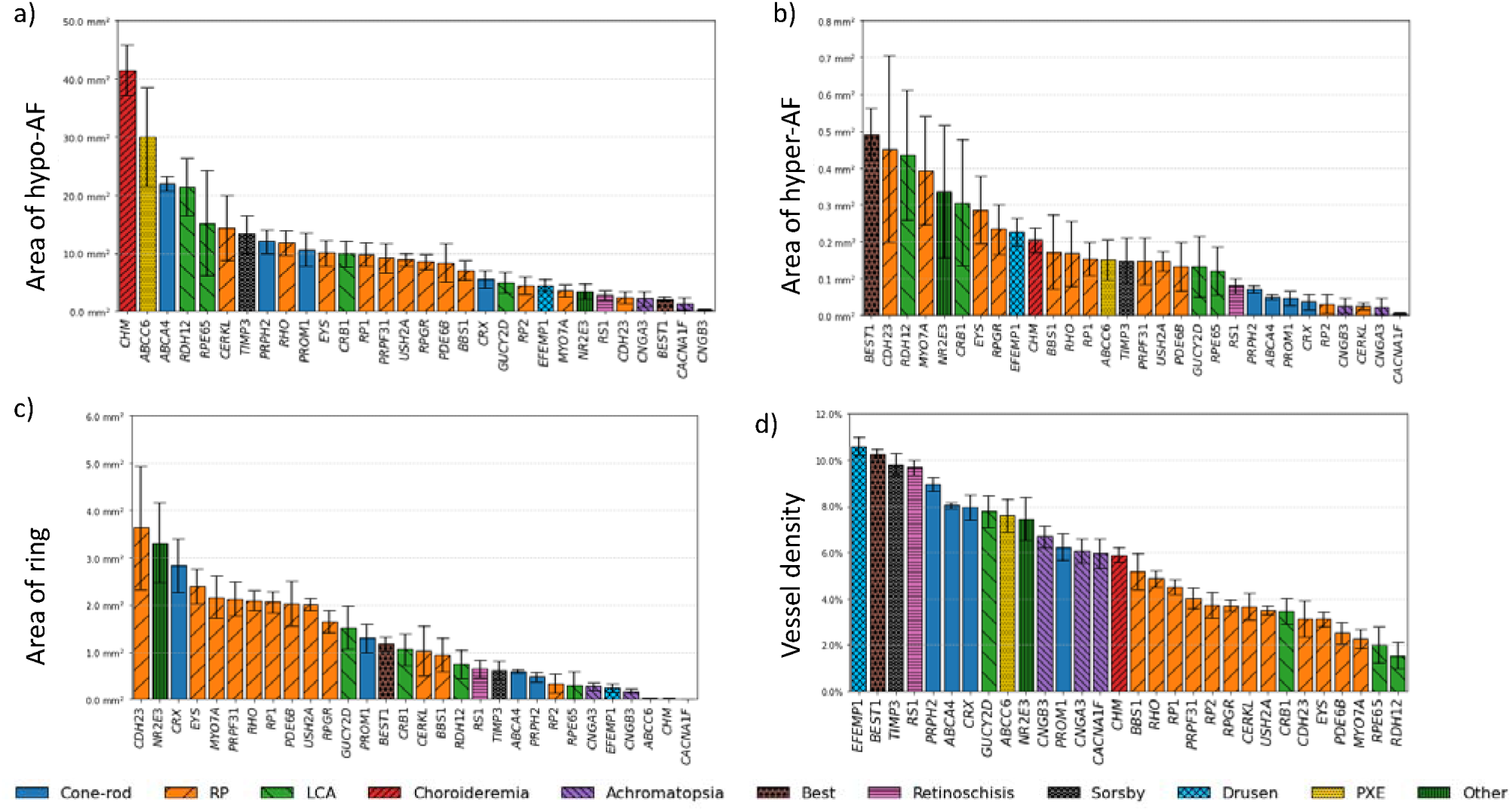
Mean **a**) extent of hypo-AF, **b**) extent of hyper-AF, **c**) extent of ring, and, **d**) Vessel density (ratio between area of vessels and total image area) across the 30 most common genes (*RPE65* included for reference). Error bars denote standard error. Values were first averaged by patient before averaging by gene to minimise correlations due to multiple contributions from individual patients. Genes are grouped into approximate phenotype groupings denoted by bar styling.

We analysed how features vary with distance from the fovea by looking at the prevalence of each feature in each 0.5 mm annulus moving away from the fovea. **Figure 3** compares prevalence of hyper- and hypo-AF at different distances from the fovea in five different genes (see **Supplementary Figure 7** for scale). The two genes associated largely with maculopathy or cone-rod dystrophy (*ABCA4*, *PRPH2*) show increased area and prevalence of hypo-AF at the fovea (**Figure 3.a** and **Figure 2.a**) but reducing proportions of the retina displaying hypo-AF moving away from the fovea. The two RP-associated genes (*USH2A*, *RPGR*) show less hypo-AF across the whole retina compared with the cone-rod genes, but with a bimodal profile, with the greatest relative proportion of hypo-AF at the fovea followed by 4-6mm from the fovea, just within the vascular arcade. For *CHM*, unlike the other genes, there was the least hypo-AF at the fovea, but substantially increased hypo-AF away from the fovea. For hyper-AF there is an increased proportion of hyper-AF at the fovea in all genes except *ABCA4* that reduces further from the fovea (**Figure 3b**). In the two RP-associated genes (*USH2A, RPGR*) there is an increase in hyper-AF at 1-3mm from the fovea.

**Figure 3:**
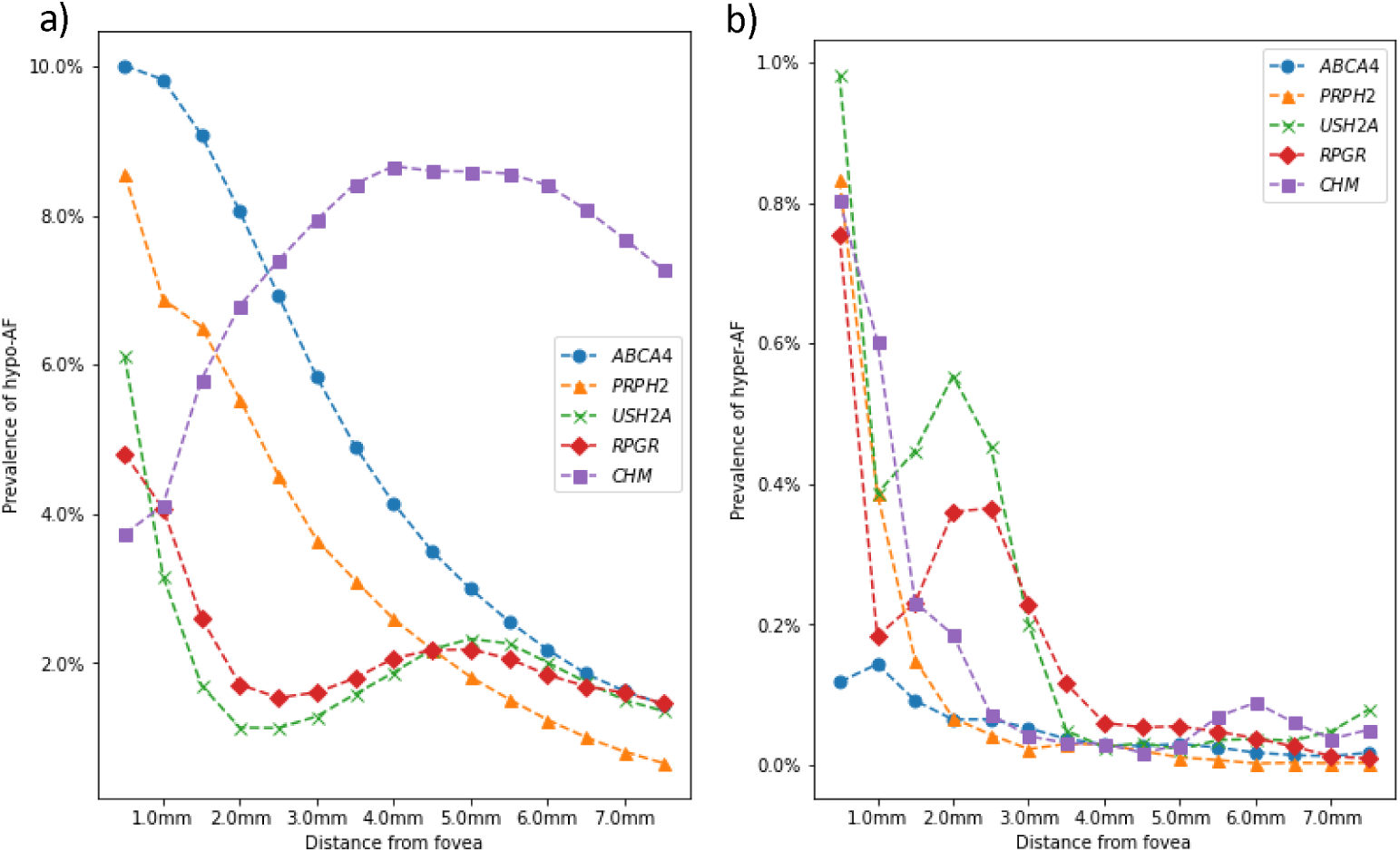
Autofluorescence (AF) as a proportion of total altered AF area in the image compared with distance from fovea for patients with variants in *ABCA4*, *RPGR*, *USH2A*, *RPGR*, and *CHM* for **a)** hypo-AF and **b)** hyper-AF.

In **Figure 4** the area of hyper-AF within 1.5mm of the fovea is compared against patient age for five different IRD genes. Most genes showed an increase with age, with the exception of *PRPH2*, which remained fairly stationary, and *BEST1*, which demonstrated a sharp decrease with patient age - although there was a considerable variability across ages within all genes.

**Figure 4:**
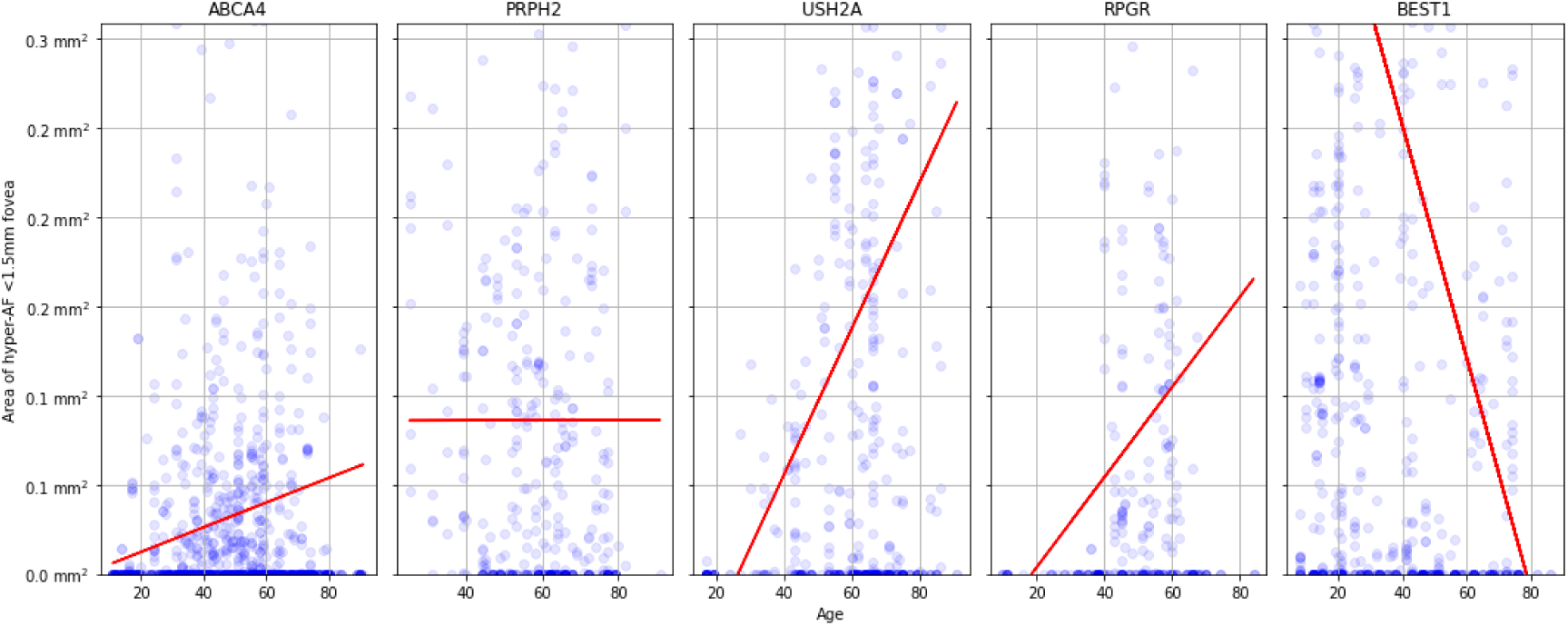
hyper-AF area within 1.5mm of the fovea (corresponding to inner 3mm ETDRs ring) compared with patient age. Least-squares regression line in red. Significant increase in hyper-AF with age for *ABCA4* (β=691 μm^2^/yr, p<0.001), *USH2A* (β=4090μm^2^/yr, p<0.001) and *RPGR* (β=2520μm^2^/yr, p<0.029). Significant decrease for *BEST1* (β=-6500μm^2^/yr, p<0.001). No significant changes of hyper-AF with age were found for *PRPH2*.

### Disease Progression

We applied AIRDetect longitudinally to monitor progression within individual patients across multiple visits. **Figure 5** shows an example using AIRDetect to visualise the decrease in ring area in individual patients with RP associated with variants in four different genes, namely *USH2A*, *PRPH2*, *RHO* and *EYS*. Comparing these four RP genes in the entire MEH IRD cohort, average rate of decrease in total ring area was greater in patients with RP associated with *EYS* (-0.178 mm^2^/year), *USH2A* (-0.066 mm^2^/year), and *RPGR* (-0.046 mm^2^/year), when compared to *RHO* (-0.040 mm^2^/year).

**Figure 5:**
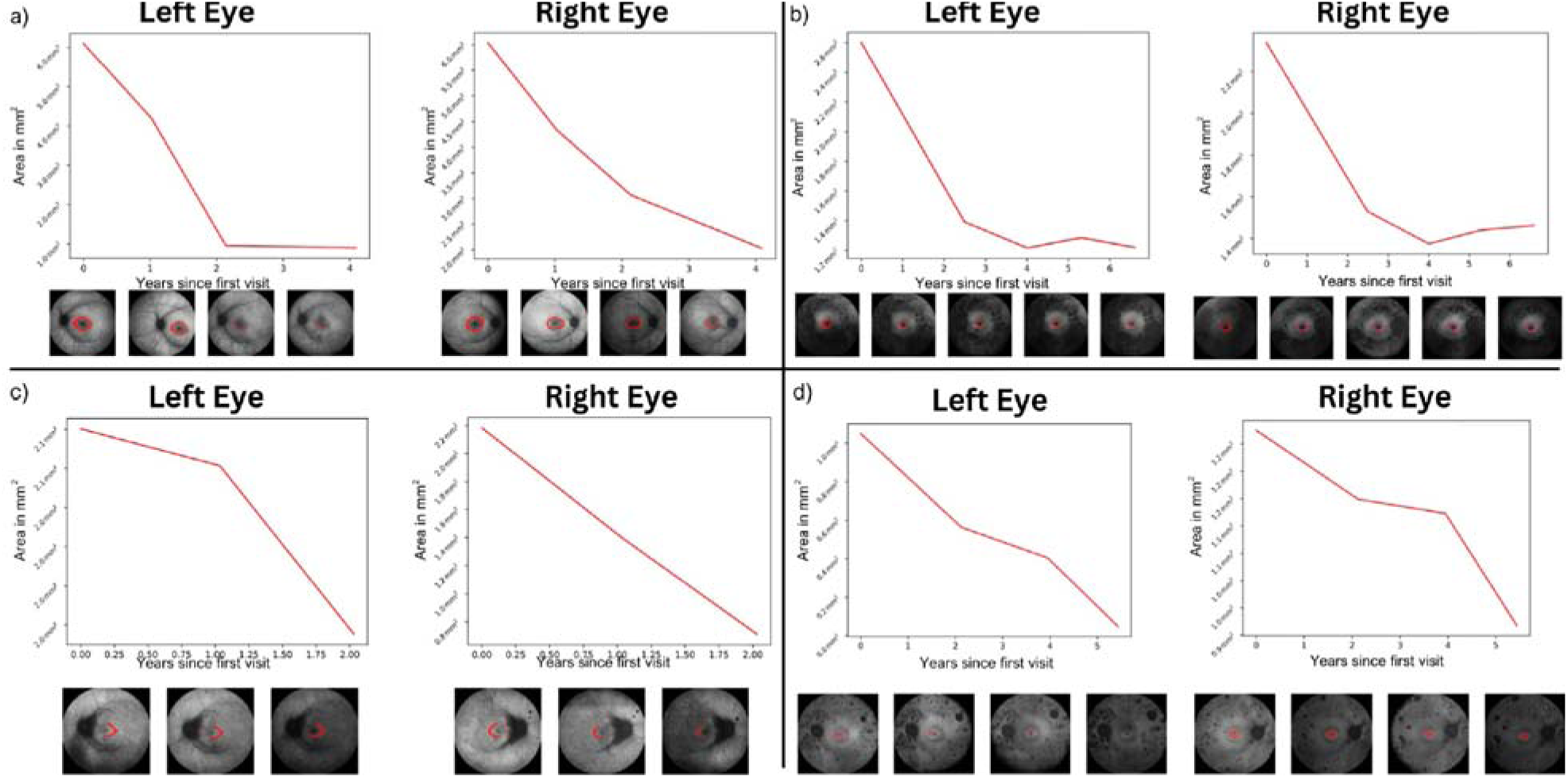
Automatic monitoring of lesion size for disease progression. Decreasing area of ring for four patients with disease-causing variants in: a) *RPGR*, b) *USH2A*, c) *RHO*, and d) *EYS*. In these genes, the macular ring is expected to shrink in diameter over time as the disease progresses.

We also applied AIRDetect to monitor progression in patients belonging to three subgroups of *ABCA4* (**Figure 6**). Patients were classified into three groups (A, B and C) based on increasing severity of genetic variants as defined by Cornelis et al. ^23,24^. Patients in group A had two severe variants, while group C had a mild variant in trans with any other variant. Patients with variants of known severity whose combination do not fit the other two groups were placed into group B. The average increase in hypo-AF area per year was compared between groups (**Supplementary Figure 3**). In keeping with previous studies ^25–29^, the mean per-patient rate of increase in hypo-AF area was highest in the highest severity classification (group A), at 3.11 mm^2^/year (0.294 mm/year change in square root area), followed by 1.59 mm^2^/year (0.169 mm/year) for the intermediate severity group (B), and finally 0.87 mm^2^/year (0.108 mm/year) in the lowest severity group (C) (**Supplementary Table 6**).

**Figure 6:**
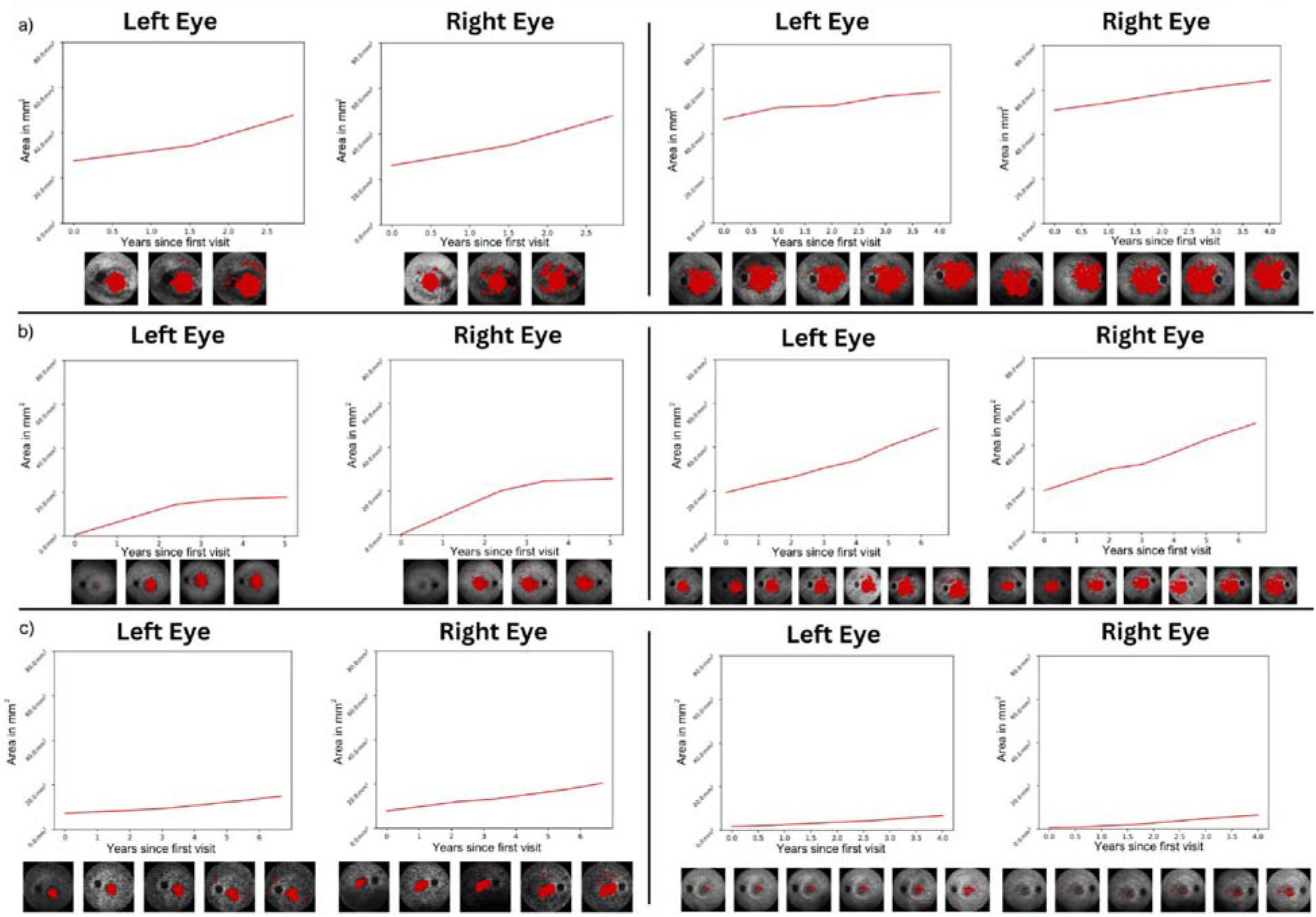
Increasing area of hypo-AF for two patients of each of the three *ABCA4* severity groups: a) group A, b) group B and c) group C. Here we see the expected patterns of progression reported in **Supplementary Table 6** with A being the fast progressors, followed by B and C.

## Discussion

The results of our cross-sectional analysis match known genotype-phenotype associations demonstrating the validity of our approach, as well yielding novel insights. For example in **Figure 2.a**, *CHM* and *ABCA4* both exhibited higher levels of hypo-AF, consistent with the large areas of atrophy that spare the fovea in choroideremia, as well as the macular atrophy typically seen in STGD1 disease (*ABCA4*) ^8,30–32^. Of interest however, *ABCC6* which is associated with pseudoxanthoma elasticum was identified to have second largest areas of hypo-AF. On further inspection, these could be explained by the large angioid streaks characteristic of this condition which can appear as hypo-AF on FAF^33^. For hyper-AF, *BEST1* exhibited the largest areas of hyper-AF, which can be attributed to the vitelliform lesion(s) that are characteristically observed in autosomal dominant and recessive forms of the disease ^34–36^ (**Figure 2.b**). For ring the presence of a macular ring typically corresponds to a demarcation between diseased and non-diseased retina, and is usually seen in RP and cone rod dystrophies, in keeping with our findings herein ^4^ (**Figure 2.c**). The lower vessel density observed in RP and LCA genes was also in keeping with the vessel attenuation commonly associated with these genes^37,38^ (**Figure 2.d**). As well as genotype-phenotype associations, we also found associations at the individual variant level confirming known association between the p.(Gly1961Glu) variant in *ABCA4* and presence of a macular ring ^39–42^ (**Supplementary Figure 3)**. When considering feature prevalence from the fovea, we found, as expected, that genes usually associated with cone-rod degeneration showed a decrease in hypo-AF extent moving away from the fovea, but with an opposite trend for the RP genes and *CHM* (**Figure 3)**. Hyper-AF was mainly concentrated at the fovea, but with a distinctive peak at 2-3mm from the fovea which may be attributed to partial macular rings classified as hyper-AF by our model (**Figure 3.b)**. *PRPH2* also had a higher coverage of hyper-AF in the fovea when compared to *ABCA4* which is consistent with the pattern/macular dystrophy and adult vitelliform phenotypes associated with *PRPH2* ^43^ (**Figure 3.b**).

In our longitudinal analysis we were able to replicate the findings of Fakin *et al.* 2016 in **Figure 7** and **Table 4**, where we found that growth of areas of hypo-AF was much more rapid in the group associated with more severe *ABCA4* genetic variants ^25^. Our estimates for rate of progression were higher than that previously reported, which may be due to the use of 55-degree as opposed to 30-degree imaging in our dataset and hence a larger area of hypo-AF ^28,44^. Comparing hyper-AF across patient age in **Figure 4**, the hyper-AF within 1.5mm of the fovea increased for *ABCA4*, *USH2A* and *RPGR*, consistent with lesions developing with disease progression over time. However, there were some noteworthy exceptions for individual genes. In particular, *BEST1* is associated with “yolk-like” regions of hyper-AF, typically within 2-3 mm of the perifovea, which change over time through pre-vitelliform, vitelliform, pseudohypopyon, vitelliruptive stages and finally to the atrophic stage ^4,36^. The highest hyper-AF signal would be associated with the vitelliform stage, progressively reducing in intensity to become a region of hypo-AF by the atrophic stage, which matches what we see as a decrease in foveal hyper-AF with age. No significant progression of hyper-AF with age was detected for *PRPH2* which is likely due to the later onset of the condition in most patients (typically after 45 years of age) and hence a more limited age range, as well as the milder pattern of dystrophy ^45^

We also identified increased rate of decrease in area of macular ring in *EYS*, *USH2A* and *RPGR* compared to *RHO* (**Figure 6**). Monitoring the rate in which the macular ring narrows down is common practice in generalised retinal dystrophies such as RP ^6^. A more rapid encroachment of the macular ring in autosomal recessive (*USH2A*, *EYS*) and X-linked (*RPGR*) genes compared to the autosomal dominant *RHO*, is consistent with the latter having a slower disease progression compared to the others ^46^.

To date, deep learning AI models to analyse FAF images from IRD patients have been limited. There have been studies developing classification models of FAF images based on IRD phenotypes ^47–50^. But as to segmentation approaches, areas of hypo-AF have been measured either manually or semi-automatically using RegionFinder on HEYEX2 software to study the progression rate of the area of atrophy in STGD1 disease ^51–54^. These approaches compared to deep-learning approaches would be challenging to scale accurately to our real-world dataset as they require considerable parameter tuning compared to deep-learning based approaches such as AIRDetect. Previous deep-learning based segmentation approaches have mostly focused on STGD1 to segment for hypo-AF^55^ or flecks^11^. Hence our AIRDetect approach represents the first to be developed and applied to a wide range of IRDs covering 170 genes.

One limitation of our approach is that the gene associations described in our study are limited by the variation in phenotypes which can occur with stage of disease for progressive conditions, different variants in the same gene or different modes of inheritance. Examining distribution of best corrected visual acuity per gene in our data, we can confirm that a range of disease stages are present in our dataset (**Supplementary Figure 8**). In terms of examples of phenotype variability per gene, *CRX* can be associated with a mild CORD but also quite severe LCA ^56–58^. *RPGR* can be associated with RP, LCA, macular dystrophy and CORD ^59,60^. We conducted a sub analysis in *ABCA4* (**Supplementary Figure 4**) but have not yet conducted this analysis across all gene variants and modes of inheritance.

Other limitations are the limited sample size for some of the genes and the large variance in imaging quality in our real-world dataset in part due to the discomfort of the patient to potential blue light-toxicity^61^, which affects the reliability of some of the features in lower quality images. While automatic image quality assessment tools exist for colour fundus retinal imaging ^62^, none have been developed for FAF imaging. Assessing image quality can also be particularly challenging for IRDs as they are associated with a wide range of pathologies, many of which can affect perceived image quality, as well as make it more challenging for the operator to acquire good quality images. We plan to develop an IRD FAF image quality assessment model in future, which should help to improve the consistency of our segmented masks and reduce noise in our analysis.

We anticipate that AIRDetect can be used to validate further clinically relevant findings, as well as identifying new potential associations between different feature patterns and certain genes or variants. Our approach could also be applied to identifying structure-function association **(Supplementary Figure 5)** as well as cross-modality image registration tasks by using vessel-based segmentation to align images (**Supplementary Figure 6**). Besides IRDs, the diverse nature of IRD-associated pathologies might make AIRDetect useful to improve robustness for segmentation of FAF imaging for other non-IRD conditions or provide a good starting point for developing models for specific conditions where data is more scarce or to other imaging modalities such as ultra-widefield imaging, via transfer learning.

In conclusion, we have conducted, to our knowledge, the largest quantitative cross-sectional and longitudinal analysis of FAF features across a diverse range of IRDs in a real world dataset, enabled by our novel automatic segmentation AI model, AIRDetect.

### Ethics

This research was approved by the IRB and the UK Health Research Authority Research Ethics Committee (REC) reference (22/WA/0049) “Eye2Gene: accelerating the diagnosis of inherited retinal diseases” Integrated Research Application System (IRAS) (project ID: 242050). All research adhered to the tenets of the Declaration of Helsinki.

### Code availability

The source code for the AIRDetect model architecture training and inference is available from https://github.com/Eye2Gene/. The model weights of AIRDetect are intellectual proprietary of UCLB so cannot be shared publicly. However, they may be shared via a licensing agreement with UCLB. A running version of the AIRDetect app is accessible via the Eye2Gene website (www.eye2gene.com) and via the Moorfields Grading Portal (grading.readingcentre.org) on invitation.

### Data availability

The data that support the findings of this study are divided into two groups, published data and restricted data. Published data are available from the Github repository. Restricted data are curated for AIRDetect users under a license and cannot be published, to protect patient privacy and intellectual property. Synthetic data derived from the test data has been made available at https://github.com/Eye2Gene/.

### Author contributions

WAW analysed the data and wrote the manuscript. NP designed the obtained the funding, designed the experiments, analysed data and wrote the manuscript. MM, KB, WAW, TACG, SAK, MDV, BM designed the experiments, analysed data and wrote the manuscript. SS analysed the data. MS wrote the manuscript. PBa analysed the data. PBu, DP analysed the data. All authors have critically reviewed the manuscript.

## Data Availability

https://www.eye2gene.com

## Data Availability

https://www.eye2gene.com

## Acknowledgement

This work is primarily funded by a NIHR AI Award (AI_AWARD02488) which supported NP, WAW, MM, KB, SD and SM. The research was also supported by a grant from the National Institute for Health Research (NIHR) Biomedical Research Centre (BRC) at Moorfields Eye Hospital NHS Foundation Trust and UCL Institute of Ophthalmology. NP was also previously funded by Moorfields Eye Charity Career Development Award (R190031A). BJ was partially funded by IIR-DE-002818 from Shire/Takeda and by the European Reference Network for Rare Malformation Syndromes, Intellectual and Other Neurodevelopmental Disorders (ERN-ITHACA). OAM is supported by the Wellcome Trust (206619/Z/17/Z). AYL is supported by an unrestricted and career development award from RPB, Latham Vision Science Awards, NIH OT2OD032644, NEI/NIH K23EY029246, and NIA/NIH U19AG066567. SA is supported by a scholarship from Qatar National Research Fund (GSRA6-1-0329-19010).This project was also supported by a generous donation by Stephen and Elizabeth Archer in memory of Marion Woods. The hardware used for analysis was supported by the BRC Challenge Fund (BRC3_027). We also gratefully acknowledge the support of NVIDIA Corporation with the donation of the Titan Xp GPU used for this research. The views expressed are those of the authors and not the funding organisations.

## Supplementary

**Supplementary Figure 1:**
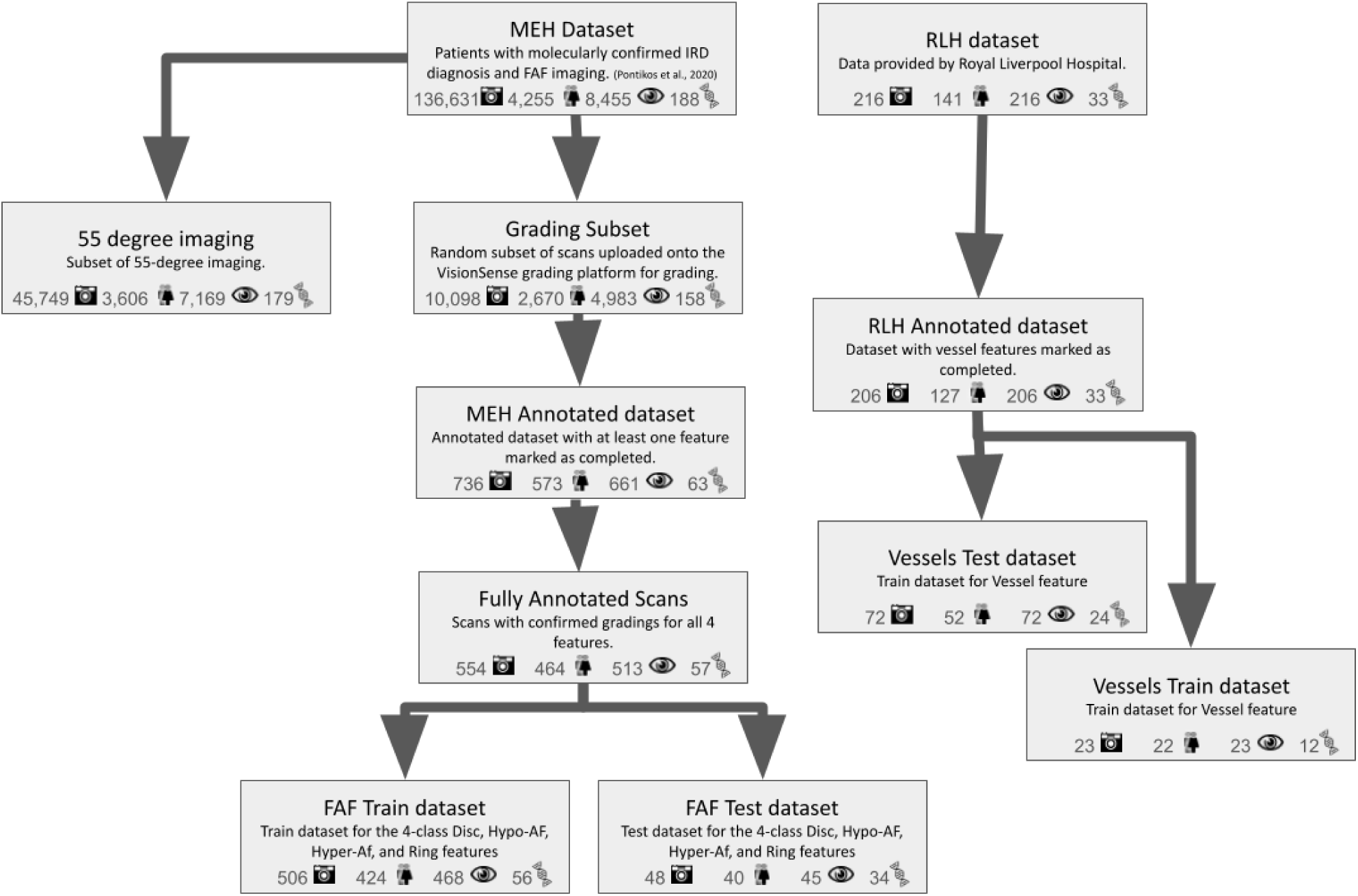
Data flowchart with number of images, patients, eyes, and genes at each stage of AIRDetect model development.

**Supplementary Figure 2:**
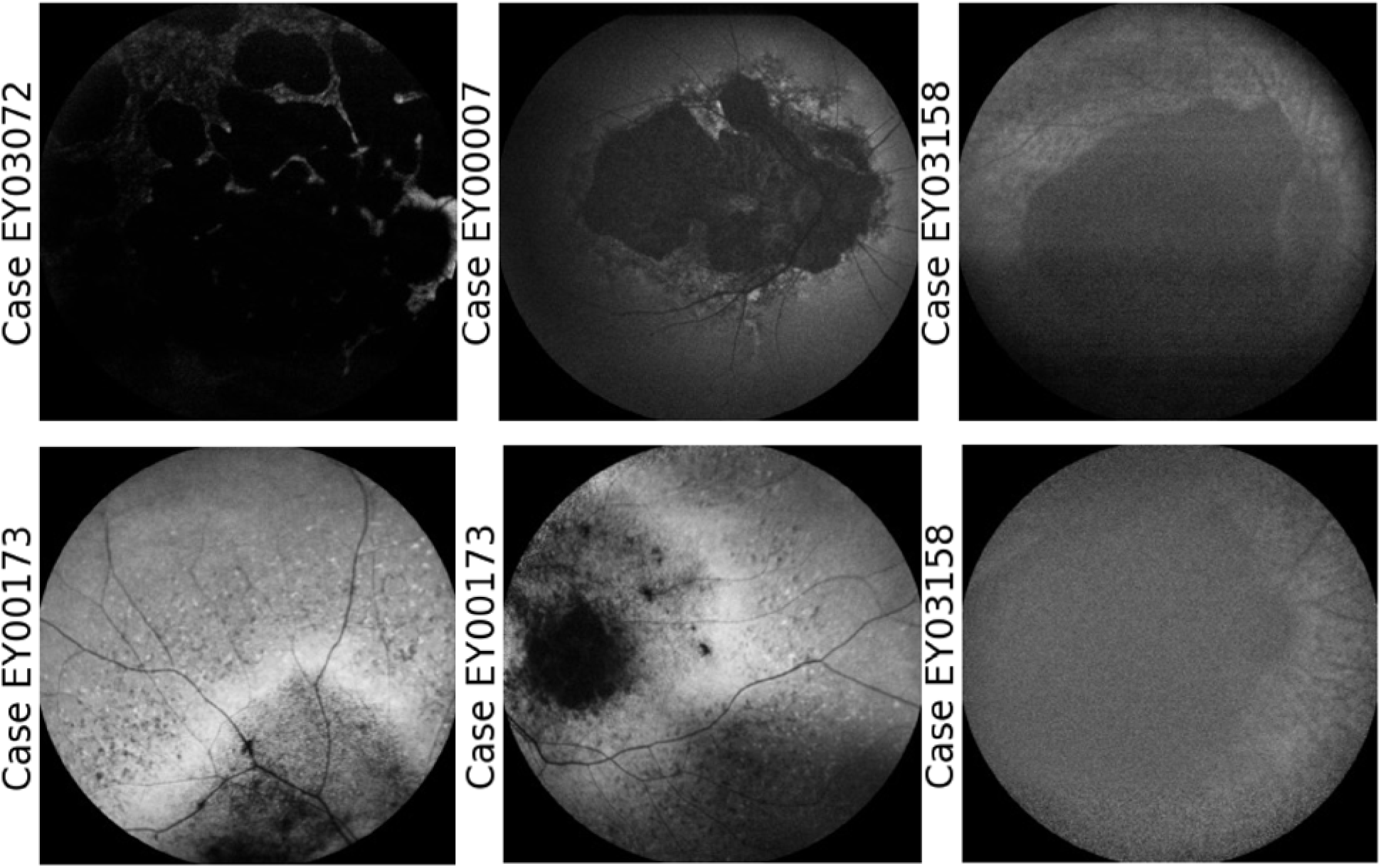
Examples of images with no Disc segmentation from the model. These are either poor quality, have significant atrophy, or are improperly centred.

**Supplementary Table 1:**
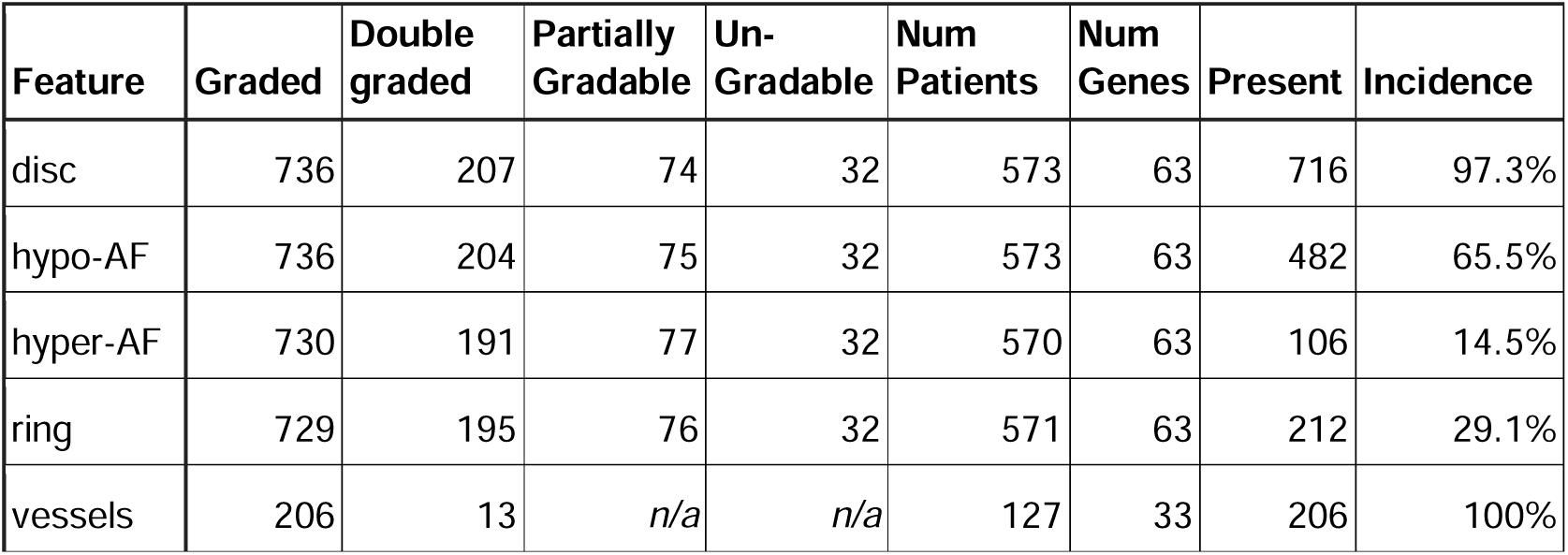
Overview of annotated dataset for the manually segmented features, considering each feature individually. Not all features were gradable within all images, with some images only annotated for some features. Images for vessel annotations were selected by clinicians and were all gradable. Incidence includes ungradable

| Feature | Graded | Double graded | Partially Gradable | Un-Gradable | Num Patients | Num Genes | Present | Incidence |
| --- | --- | --- | --- | --- | --- | --- | --- | --- |
| disc | 736 | 207 | 74 | 32 | 573 | 63 | 716 | 97.3% |
| hypo-AF | 736 | 204 | 75 | 32 | 573 | 63 | 482 | 65.5% |
| hyper-AF | 730 | 191 | 77 | 32 | 570 | 63 | 106 | 14.5% |
| ring | 729 | 195 | 76 | 32 | 571 | 63 | 212 | 29.1% |
| vessels | 206 | 13 | *n/a* | *n/a* | 127 | 33 | 206 | 100% |

**Supplementary Table 2:** Vessel metrics and their description.

| Vessel Metric | Description |
| --- | --- |
| Fractal Dimension | Method that represents geometric complexity of the vascular branching pattern observed in the retina. Essentially how close are the vessels to being “space-filling”. |
| Vessel Density | Ratio between area of vessels and total image area. |
| Average Width | Average width of vessels. |
| Distance Tortuosity | Distance tortuosity is a measure of the tortuosity of a path based on the ratio of the actual path length to the straight-line distance between the start and end points of the path <sup>63</sup> |
| Squared Curvature Tortuosity | Squared curvature tortuosity is a more sophisticated measure of tortuosity that takes into account the curvature along the path, providing a detailed view of its winding nature <sup>63</sup> |
| Tortuosity Density | Assesses vessel tortuosity by aggregating local contributions, examining the degree to which each turn curve deviates from a smooth curve <sup>64</sup> |

**Supplementary Table 3:** Feature statistics by gene for selected 30 genes. Results are mean across all images. %=Incidence, A=Average Area (mm^2^), C=Num Components, I = Intensity (pixel brightness), <3mm=Proportion of feature area within 3mm of the fovea (corresponding to outer 6mm ETRDS ring), D=vessel density, F=fractal dimension. The table cells have been shaded with lower values in red, intermediate values in white and larger values in green.

| Gene | disc |  |  |  | hypo-AF |  |  |  |  | hyper-AF |  |  |  |  | ring |  |  |  |  | vessels |  |
| --- | --- | --- | --- | --- | --- | --- | --- | --- | --- | --- | --- | --- | --- | --- | --- | --- | --- | --- | --- | --- | --- |
|  | A | C | I | <3mm | % | A | C | I | <3mm | % | A | C | I | <3mm | % | A | C | I | <3mm | D | F |
| ABCA4 | 2.19 | 1.01 | 9.20% | 13.70% | 80.30% | 19.81 | 1.45 | 11.10% | 61.30% | 18.00% | 0.05 | 0.33 | 66.40% | 48.30% | 33.50% | 0.57 | 1.58 | 59.60% | 81.80% | 7.65% | 1.33 |
| ABCC6 | 1.53 | 1.03 | 16.90% | 17.60% | 69.00% | 21.44 | 2.08 | 14.30% | 32.60% | 54.90% | 0.23 | 1.2 | 62.50% | 23.40% | 8.20% | 0.03 | 0.15 | 43.30% | 38.00% | 8.07% | 1.37 |
| BBS1 | 2.59 | 1.02 | 10.40% | 7.50% | 67.50% | 8.35 | 1.29 | 11.90% | 56.90% | 25.90% | 0.15 | 0.38 | 47.30% | 86.90% | 32.50% | 0.84 | 1.03 | 56.50% | 72.70% | 4.56% | 1.18 |
| BEST1 | 1.84 | 1.01 | 11.60% | 4.10% | 40.50% | 2.58 | 0.9 | 11.50% | 59.70% | 57.50% | 0.73 | 1.09 | 61.20% | 59.80% | 57.50% | 1.28 | 1.83 | 56.30% | 77.20% | 9.62% | 1.38 |
| CACNA1F | 2.04 | 1.01 | 14.80% | 0.00% | 5.70% | 0.91 | 0.2 | 7.10% | 42.40% | 12.80% | 0.01 | 0.15 | 59.80% | 2.50% | 3.80% | 0.01 | 0.06 | 53.60% | 79.80% | 5.35% | 1.21 |
| CDH23 | 2.05 | 1.01 | 14.30% | 6.40% | 16.20% | 0.71 | 0.47 | 10.40% | 42.00% | 19.60% | 0.23 | 0.5 | 57.60% | 34.20% | 77.30% | 2.46 | 2.73 | 47.30% | 67.30% | 2.46% | 1.05 |
| CERKL | 2.63 | 1 | 12.30% | 9.10% | 82.30% | 10.33 | 1.43 | 12.10% | 51.70% | 17.70% | 0.02 | 0.21 | 46.00% | 88.60% | 29.70% | 1.04 | 0.95 | 58.10% | 65.30% | 3.51% | 1.16 |
| CHM | 1.45 | 1.06 | 20.60% | 2.80% | 82.60% | 51.77 | 3.28 | 19.90% | 21.00% | 48.80% | 0.26 | 0.83 | 54.50% | 37.90% | 5.50% | 0.03 | 0.08 | 43.20% | 32.20% | 6.20% | 1.3 |
| CNGA3 | 1.74 | 1.01 | 13.80% | 14.50% | 31.10% | 1.48 | 0.96 | 7.10% | 56.30% | 8.00% | 0.05 | 0.17 | 54.00% | 31.80% | 35.60% | 0.32 | 0.75 | 50.00% | 74.90% | 5.36% | 1.24 |
| CNGB3 | 1.91 | 0.99 | 14.90% | 12.70% | 12.50% | 0.16 | 0.38 | 6.70% | 73.60% | 10.40% | 0.01 | 0.12 | 52.70% | 63.90% | 32.60% | 0.2 | 0.63 | 54.60% | 92.20% | 5.92% | 1.24 |
| CRB1 | 1.97 | 1.02 | 14.70% | 3.60% | 55.20% | 8.3 | 1.22 | 15.60% | 40.40% | 44.00% | 0.41 | 0.89 | 46.40% | 54.00% | 43.70% | 1.73 | 1.35 | 48.90% | 59.20% | 4.27% | 1.11 |
| CRX | 2.11 | 1 | 15.60% | 5.70% | 70.30% | 5.51 | 1.39 | 15.60% | 62.70% | 16.90% | 0.05 | 0.26 | 50.70% | 29.90% | 59.30% | 2.23 | 4.22 | 52.90% | 65.90% | 7.30% | 1.33 |
| EFEMP1 | 1.77 | 1 | 15.30% | 6.00% | 78.70% | 5.15 | 1.84 | 14.20% | 82.40% | 77.90% | 0.24 | 1.55 | 61.80% | 79.40% | 24.10% | 0.18 | 0.66 | 60.00% | 82.50% | 10.10% | 1.43 |
| EYS | 2.36 | 1.02 | 15.20% | 2.90% | 65.90% | 9.55 | 1.64 | 13.30% | 27.30% | 23.40% | 0.23 | 0.4 | 57.60% | 42.90% | 72.50% | 2.08 | 2.76 | 52.60% | 76.80% | 3.07% | 1.16 |
| GUCY2D | 2 | 1 | 14.70% | 6.50% | 48.00% | 5.25 | 0.86 | 17.50% | 71.20% | 17.60% | 0.22 | 0.3 | 61.20% | 36.00% | 60.90% | 1.51 | 2.94 | 53.10% | 75.00% | 6.91% | 1.29 |
| MYO7A | 2.07 | 0.99 | 15.80% | 2.30% | 42.30% | 2.37 | 0.95 | 18.00% | 32.90% | 28.00% | 0.41 | 0.97 | 53.70% | 21.80% | 73.60% | 3.24 | 3.26 | 47.00% | 57.80% | 2.70% | 1.09 |
| NR2E3 | 1.94 | 1.02 | 10.90% | 7.40% | 31.50% | 1.42 | 0.76 | 10.20% | 12.90% | 21.40% | 0.37 | 0.5 | 56.50% | 17.90% | 39.60% | 1.72 | 1.4 | 52.20% | 24.70% | 8.33% | 1.32 |
| PDE6B | 2.62 | 1.01 | 14.80% | 3.40% | 52.20% | 4.48 | 0.93 | 17.00% | 49.30% | 21.30% | 0.08 | 0.28 | 55.10% | 56.90% | 86.00% | 3.37 | 4.06 | 47.30% | 86.60% | 3.00% | 1.12 |
| PROM1 | 2.3 | 1.01 | 12.60% | 7.00% | 80.00% | 12.15 | 1.72 | 12.60% | 72.00% | 21.10% | 0.04 | 0.25 | 52.30% | 72.90% | 34.80% | 0.85 | 2.06 | 54.00% | 82.50% | 6.10% | 1.26 |
| PRPF31 | 2.2 | 1.03 | 15.50% | 1.20% | 48.10% | 6.78 | 1.56 | 12.00% | 41.30% | 18.80% | 0.12 | 0.34 | 45.90% | 68.30% | 64.00% | 2.17 | 2.85 | 52.50% | 86.90% | 4.10% | 1.21 |
| PRPH2 | 2.05 | 1.01 | 12.30% | 4.50% | 68.70% | 10.28 | 1.89 | 10.10% | 62.00% | 36.40% | 0.07 | 0.58 | 59.10% | 77.10% | 26.30% | 0.69 | 0.85 | 55.80% | 69.40% | 8.00% | 1.36 |
| RDH12 | 1.92 | 1.01 | 18.00% | 11.80% | 58.10% | 22.35 | 1.1 | 19.00% | 37.30% | 33.70% | 0.37 | 0.52 | 49.00% | 22.10% | 34.10% | 1.51 | 2.52 | 57.90% | 84.80% | 2.93% | 0.99 |
| RHO | 2.17 | 1.03 | 15.50% | 1.50% | 61.90% | 9.74 | 1.72 | 13.60% | 34.80% | 18.50% | 0.11 | 0.24 | 53.00% | 56.00% | 70.60% | 2.25 | 2.99 | 53.60% | 82.80% | 4.47% | 1.22 |
| RP1 | 2.19 | 1.01 | 13.70% | 5.50% | 63.00% | 9.32 | 1.76 | 13.10% | 40.10% | 19.10% | 0.18 | 0.27 | 55.90% | 66.70% | 65.00% | 2.03 | 2.98 | 54.60% | 74.80% | 4.59% | 1.23 |
| RP2 | 2.48 | 1.05 | 9.90% | 13.30% | 47.60% | 3.1 | 1.19 | 10.50% | 48.30% | 6.90% | 0.02 | 0.08 | 54.70% | 84.40% | 13.00% | 0.26 | 0.38 | 59.00% | 67.80% | 3.65% | 1.19 |
| RPE65 | 1.15 | 1.06 | 19.30% | 7.60% | 52.40% | 40.13 | 2.65 | 15.00% | 29.10% | 37.80% | 0.24 | 0.65 | 56.40% | 61.80% | 4.90% | 0.19 | 0.05 | 43.10% | 97.60% | 2.04% | 0.97 |
| RPGR | 2.31 | 1.01 | 13.70% | 3.70% | 54.20% | 7.78 | 1.27 | 14.40% | 36.50% | 22.60% | 0.13 | 0.31 | 54.40% | 58.60% | 61.30% | 2.13 | 2.09 | 55.90% | 82.00% | 4.06% | 1.19 |
| RS1 | 1.93 | 1 | 11.90% | 3.50% | 26.80% | 2.41 | 0.58 | 10.70% | 36.70% | 24.80% | 0.1 | 0.37 | 44.30% | 73.20% | 16.90% | 0.35 | 0.59 | 53.00% | 66.40% | 8.69% | 1.34 |
| TIMP3 | 1.71 | 1.01 | 17.50% | 9.50% | 62.10% | 16.35 | 1.84 | 15.30% | 46.50% | 30.70% | 0.14 | 0.51 | 58.50% | 32.60% | 27.00% | 0.47 | 1 | 55.20% | 35.30% | 9.15% | 1.39 |
| USH2A | 2.17 | 1.02 | 15.60% | 2.30% | 61.30% | 8.47 | 1.76 | 13.00% | 36.50% | 19.80% | 0.15 | 0.31 | 53.50% | 66.50% | 72.10% | 1.98 | 3.2 | 53.50% | 87.50% | 3.42% | 1.17 |
| All | 2.09 | 1.02 | 13.50% | 6.90% | 61.20% | 13.04 | 1.46 | 13.00% | 48.20% | 25.10% | 0.15 | 0.43 | 57.10% | 52.80% | 43.70% | 1.19 | 1.78 | 54.10% | 77.90% | 5.75% | 1.25 |

**Supplementary Table 4: Feature statistics for all genes.** Phenotypes: pheno = most common phenotype presentation according to literature. ACHM = achromatopsia, ALB = albinism, BEST = best disease, CD = cone-dystrophy, CHM = choroidemia, CR = cone-rod, CSNB = congenital stationary night blindness, DR = diabetic retinopathy, FEVR = Familial exudative vitreoretinopathy, GA = Gyrate atrophy, LCA = Leber’s congenital amaurosis, MAC = Microphthalmia, anophthalmia, coloboma, MD = macular dystrophy, OA = optic atrophy, PD = pattern dystrophy, PXE = pseudoxanthoma elasticum, RP = retinitis pigmentosa. pat = number of patients. img = number of FAF images. Feature metrics are averaged across all images per gene. Features: % = average incidence in percent, A = averge area in mm^2^, C = average number of clusters, I = average pixel intensity in percentage, % <6mm = average incidence within 6mm area in percent, FD = vessel fractal dimensions, D = average vessel density, W = average vessel width, DTM = distance tortuosity mean, SCTM = squared curvature tortuosity mean, TDM = tortuosity density mean. (see online supplementary table 4)

**Supplementary Table 5:** All vessel metrics across selected genes. Definitions of metrics are given in **Supplementary Table 2**. The table cells have been shaded with lower values in red, intermediate values in white and larger values in green.

| gene | Fractal Dimension | Vessel Density | Average Width | Distance Tortuosity Mean | Squared Curvature Tortuosity Mean | Tortuosity Density Mean |
| --- | --- | --- | --- | --- | --- | --- |
| *ABCA4* | 1.33 | 7.65% | 184.81 | 4.81 | 61.6 | 0.71 |
| *ABCC6* | 1.37 | 8.07% | 196.68 | 6.17 | 80.7 | 0.73 |
| *BBS1* | 1.18 | 4.56% | 186.86 | 9.66 | 214.9 | 0.7 |
| *BEST1* | 1.38 | 9.62% | 203.18 | 4.46 | 49.4 | 0.71 |
| *CACNA1F* | 1.21 | 5.35% | 218.03 | 9.24 | 83.8 | 0.73 |
| *CDH23* | 1.05 | 2.46% | 198.91 | 7.97 | 177.8 | 0.68 |
| *CERKL* | 1.16 | 3.51% | 179.55 | 7.2 | 95.8 | 0.67 |
| *CHM* | 1.3 | 6.20% | 190.34 | 6.14 | 104.4 | 0.69 |
| *CNGA3* | 1.24 | 5.36% | 209.19 | 5.17 | 70.9 | 0.7 |
| *CNGB3* | 1.24 | 5.92% | 205.53 | 6.42 | 67.1 | 0.7 |
| *CRB1* | 1.11 | 4.27% | 178.82 | 7.2 | 83.2 | 0.69 |
| *CRX* | 1.33 | 7.30% | 192.4 | 9.58 | 119.5 | 0.72 |
| *EFEMP1* | 1.43 | 10.06% | 208.55 | 4.31 | 39.4 | 0.72 |
| *EYS* | 1.16 | 3.07% | 172.31 | 7.39 | 108.3 | 0.69 |
| *GUCY2D* | 1.29 | 6.91% | 197.87 | 5.41 | 64.4 | 0.72 |
| *MYO7A* | 1.09 | 2.70% | 182.09 | 11 | 296.8 | 0.7 |
| *NR2E3* | 1.32 | 8.33% | 202.83 | 5.85 | 77.5 | 0.72 |
| *PDE6B* | 1.12 | 3.00% | 181.7 | 7.22 | 113.4 | 0.71 |
| *PROM1* | 1.26 | 6.10% | 179.6 | 6.5 | 116.9 | 0.72 |
| *PRPF31* | 1.21 | 4.10% | 178.26 | 7.11 | 146.2 | 0.7 |
| *PRPH2* | 1.36 | 8.00% | 184.62 | 4.76 | 52.1 | 0.71 |
| *RDH12* | 0.99 | 2.93% | 179.48 | 3.48 | 20.6 | 0.72 |
| *RHO* | 1.22 | 4.47% | 175.72 | 6.49 | 86.1 | 0.69 |
| *RP1* | 1.23 | 4.59% | 174.37 | 6.93 | 139.4 | 0.7 |
| *RP2* | 1.19 | 3.65% | 181.19 | 6.83 | 109.3 | 0.69 |
| *RPE65* | 0.97 | 2.04% | 164.35 | 6.64 | 110.8 | 0.65 |
| *RPGR* | 1.19 | 4.06% | 182.93 | 8.03 | 126.1 | 0.7 |
| *RS1* | 1.34 | 8.69% | 207.5 | 5.45 | 62.3 | 0.72 |
| *TIMP3* | 1.39 | 9.15% | 198.27 | 4.99 | 71.9 | 0.71 |
| *USH2A* | 1.17 | 3.42% | 170.22 | 7.26 | 101.6 | 0.69 |
| **All** | **1.25** | **5.75%** | **186.52** | **6.45** | **98** | **0.7** |

**Supplementary Table 6:**
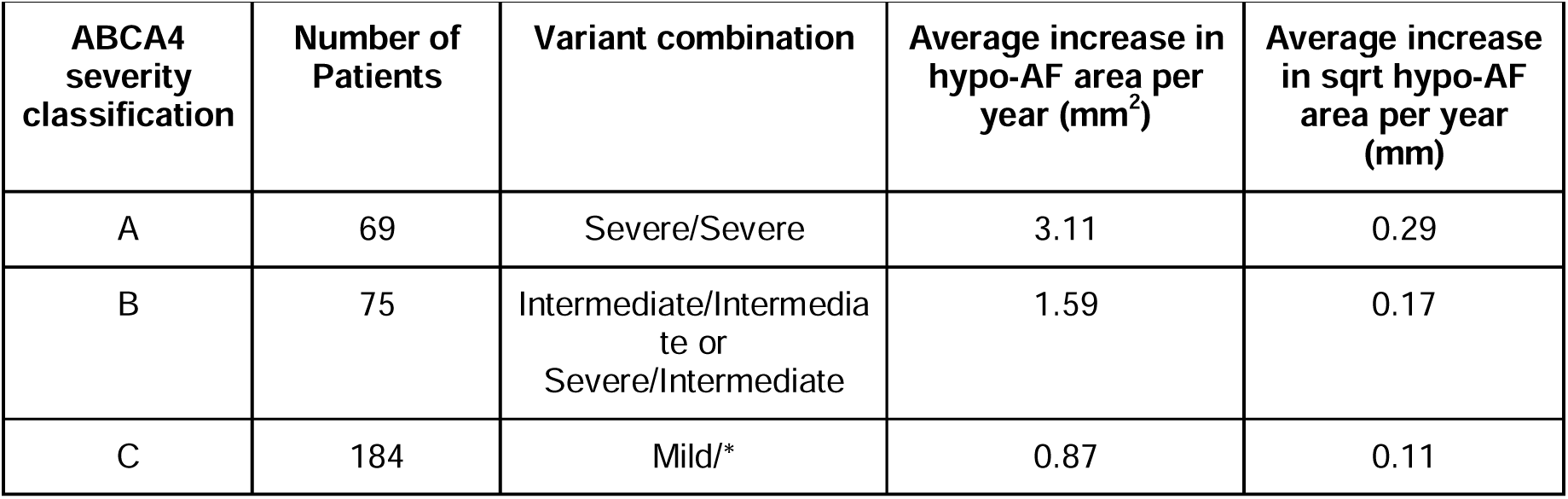
Average increase in hypo-AF area stratified by ABCA4 variant severity. ABCA4 patients are grouped based on the severity of their genetic variants as proposed by Cornelis et al. 2022 into groups A, B and C ^23^.

**Supplementary Figure 3:**
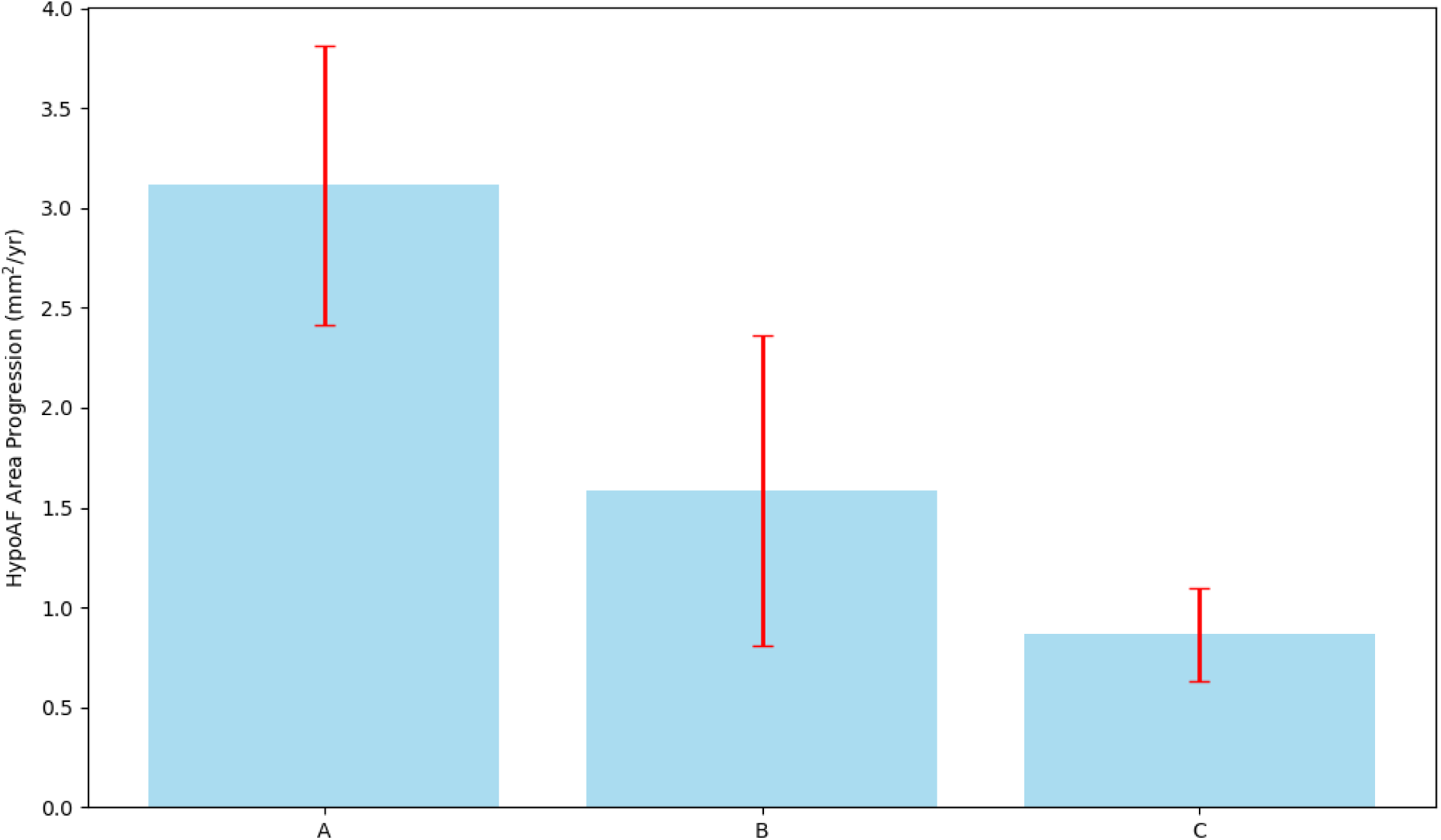
Rate of progression of hypo-AF in mm^2^ per year for patients in the three severity classification groups of *ABCA4*. Note that Group A has a higher mean rate of progression than groups B and C, as it corresponds to the group with the highest severity. Error bars denote standard error.

**Supplementary Figure 4:**
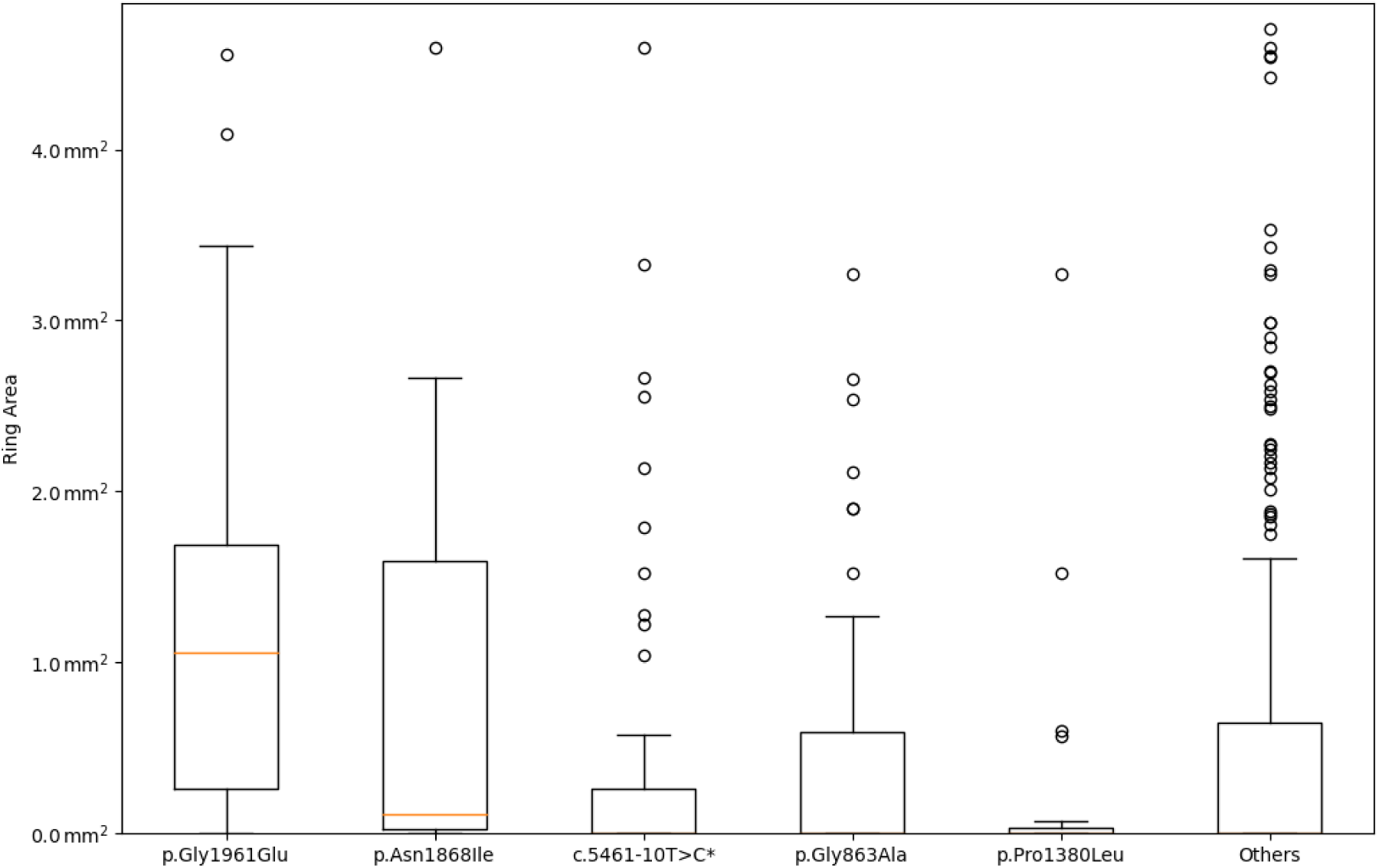
Comparison of the mean per-patient extent of macular ring present for patients with different variants (i.e. patients with at least one copy of the given variant) in *ABCA4*. Axes are truncated to exclude 99th percentile outliers. Most variants of *ABCA4* are not associated with a macular ring of raised AF, apart from p.(Gly1961Glu) which we see reflected in the different distributions of ring area in our data.

**Supplementary Figure 5:**
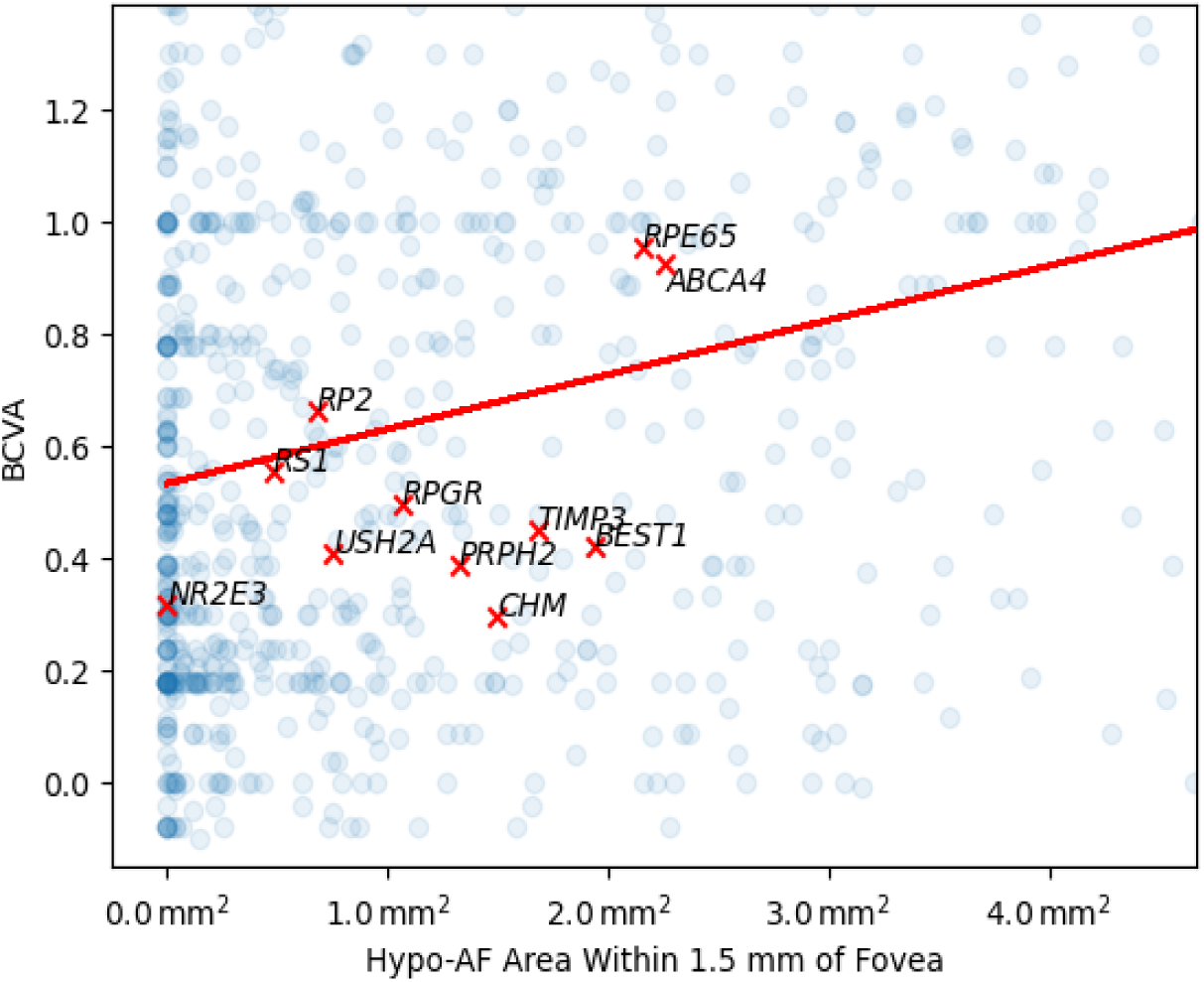
hypo-AF area within 1.5mm of the fovea compared to LogMAR best corrected visual acuity (BCVA) where higher values corresponds to poorer acuity. Axes rescaled to 90th pct of data for legibility. Each circle represents a single patient with mean value across images. Least-squares regression line in red (β=0.083, p<0.001). Mean values for select genes are indicated by red crosses. Comparing hypo-AF area within 1.5mm of the fovea and LogMAR best corrected visual acuity (BCVA) showed a positive statistical association (β=0.083, p<0.001). However, some genes demonstrated a different relationship from the main trend. For example, in *ABCA4* a worse BCVA was observed than might be expected from hypo-AF coverage, likely because *ABCA4*-associated retinopathy usually initially affects the fovea/central macula. By contrast, *CHM* typically exhibits a spared foveal island despite having significant areas of atrophy, thus accounting for the relatively preserved BCVA.

**Supplementary Figure 6:**
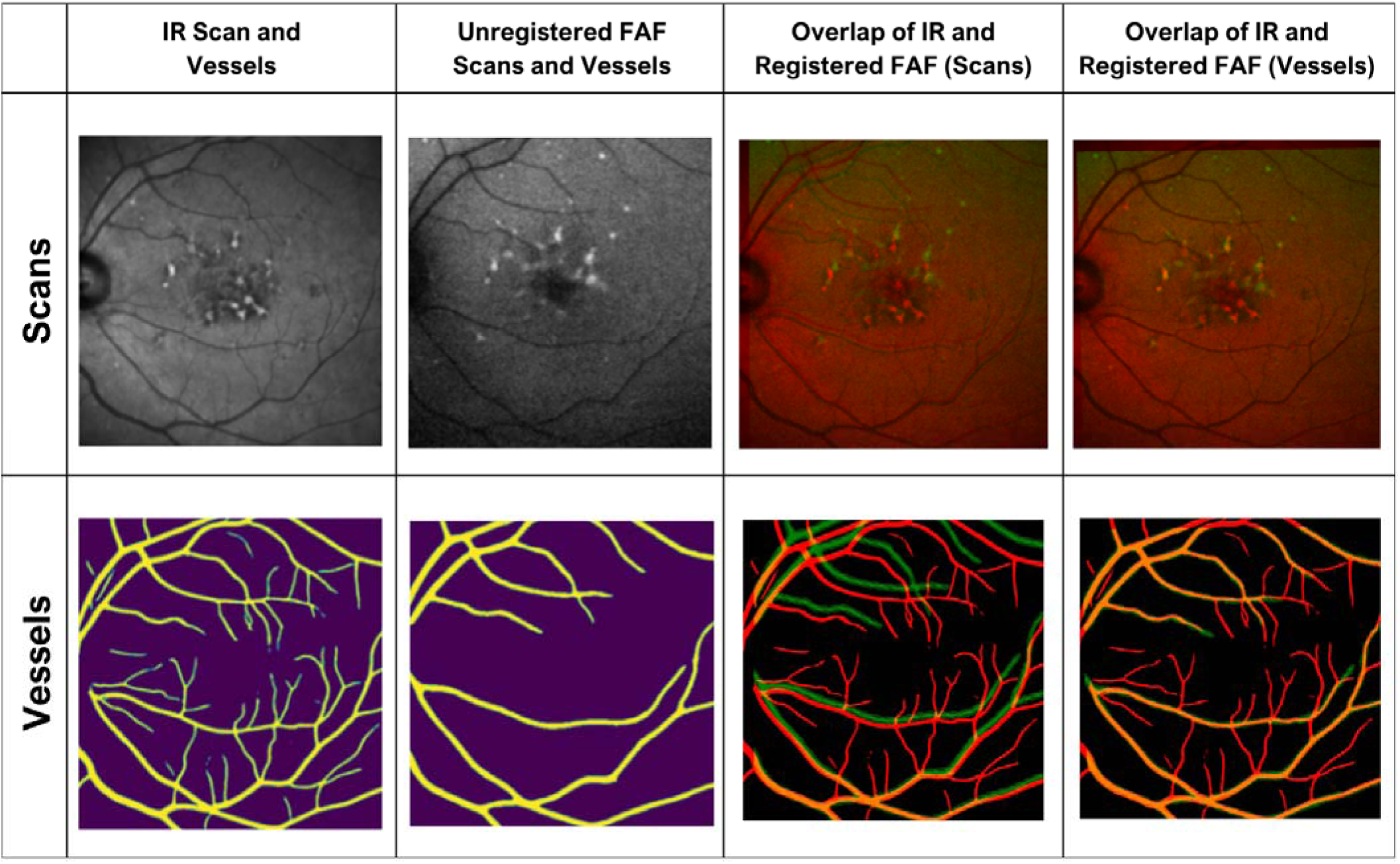
Example showing how vessel tree segmentation improves cross-modality image registration. First row shows the individual and overlaid images, and second rows shows corresponding segmented vessel masks. For the overlaid images, the IR image is rendered in red, while the FAF image is rendered in green, enabling overlap to be assessed by looking at the correspondence between the two-colour channels. Vessel trees were extracted using AIRDetect for both the IR and the FAF image. Results of automatic registration directly on the raw images (scans column) and registration on the vessel trees (vessels column) are shown. In both cases this registration was performed using the SimpleElastix package. As shown by the final column, registering using vessel trees results in better overlap than registering using images alone.

**Supplementary Figure 7:**
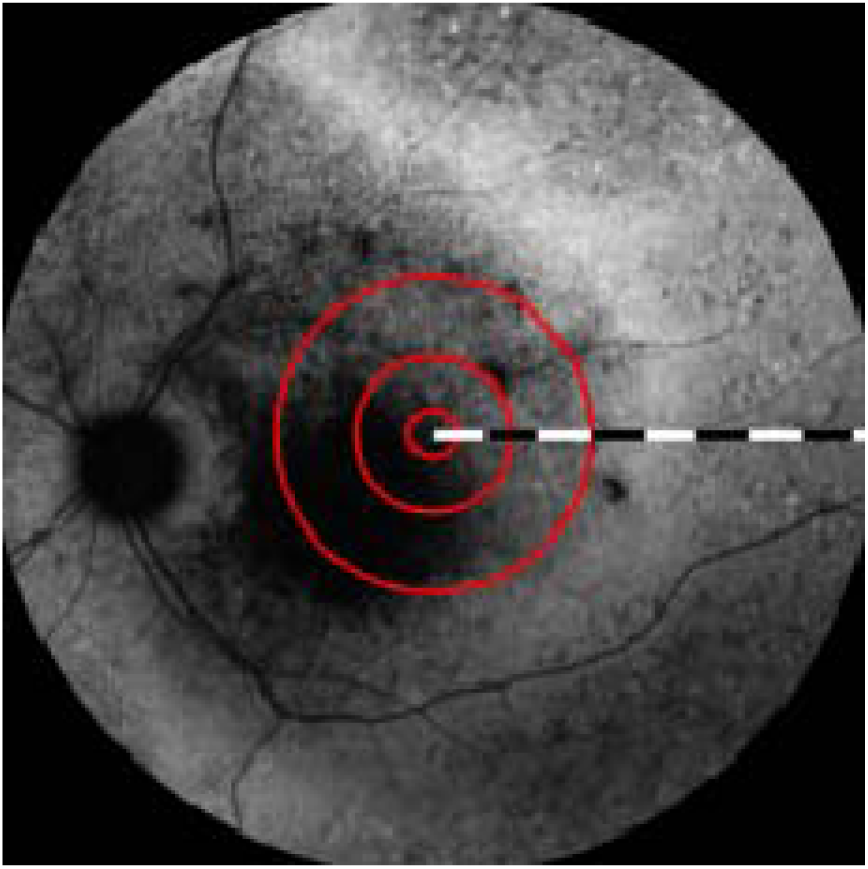
55-degree FAF image with 0.5mm, 1.5mm, and 3mm radial distances shown (corresponding to 1mm, 3mm, and 6mm diameter ETDRS regions), and scale bar with 1mm gradations.

**Supplementary Figure 8:**
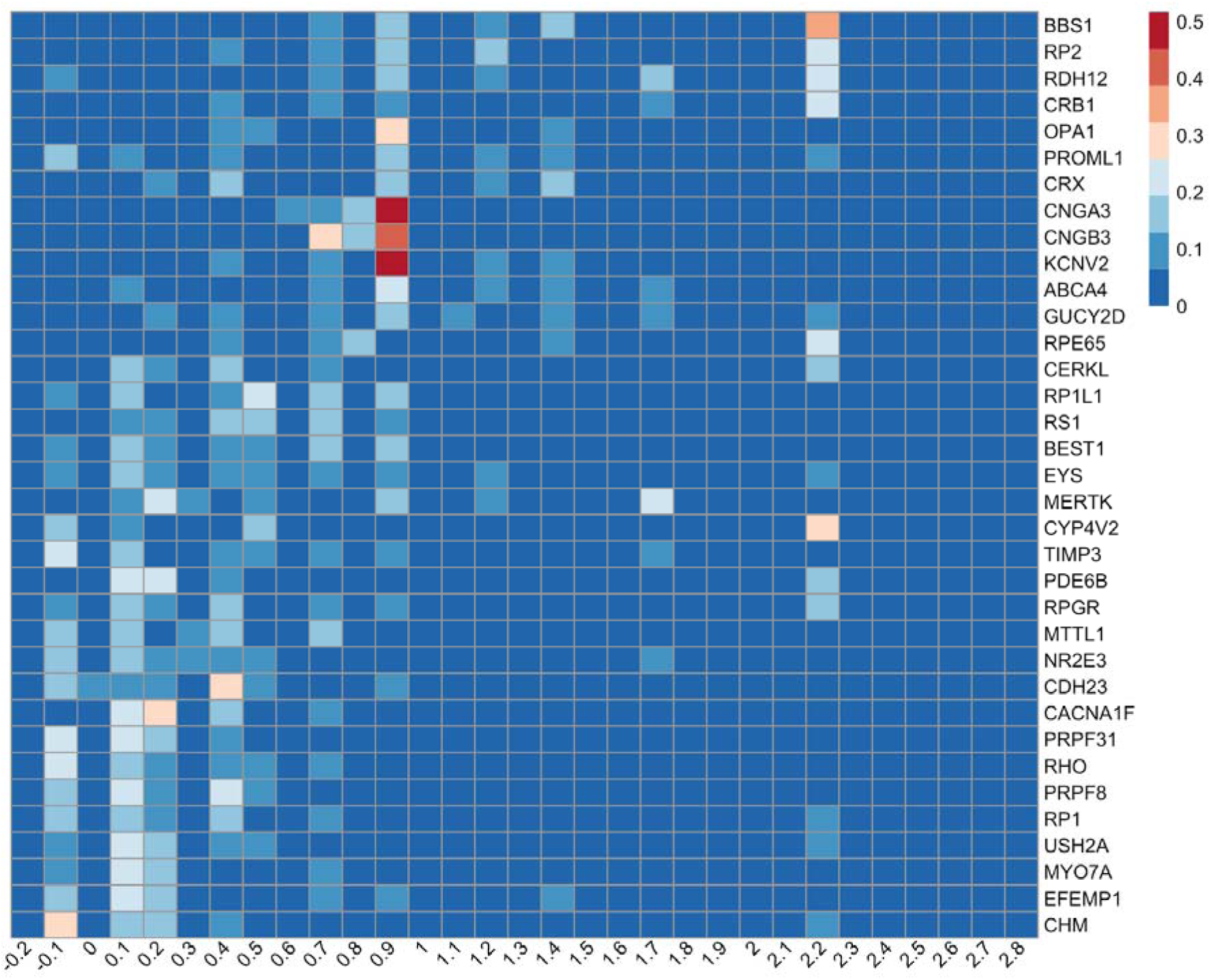
Distributions of best corrected visual acuity (in LogMar) with respect to 36 genes sorted by the median of the visual acuity distribution per gene. Low vision is defined as a best-corrected visual acuity worse than 0.5 LogMAR but equal or better than 1.3 LogMAR in the better eye. Blindness is defined as a best-corrected visual acuity worse than 1.3 LogMAR. Also represented are Logmar of 1.98 (Counting Fingers), 2.28 (Hand Movement) and 2.7 (Light Perception).

