## Supplementary Table 4 for "Quantification of Fundus Autofluorescence Features in a Molecularly Characterized Cohort of More Than 3500 Inherited Retinal Disease Patients from the United Kingdom"

| | | | | | | Disc | | | | Hypo-AF | | | | | Hyper-AF | | | | | Ring | | | | | Vessels | | | | | |
|---|---|---|---|---|---|---|---|---|---|---|---|---|---|---|---|---|---|---|---|---|---|---|---|---|---|---|---|---|---|---|
| gene | patient count | image count | Patient Ages | | | area | num clusters | intensity | % <6mm | incidence | area | num clusters | intensity | % <6mm | incidence | area | num clusters | intensity | % <6mm | incidence | area | num clusters | intensity | % <6mm | Fractal Dimension | Density | Width | Distance tortuosity mean | Squared curvature tortuosity mean | Tortuosity density mean |
| **Total** | **3491** | **32964** | **min** | **median** | **max** | **2.09** | **1.02** | **13.51%** | **6.86%** | **61.15%** | **13.04** | **1.46** | **13.02%** | **48.17%** | **25.11%** | **0.15** | **0.43** | **57.14%** | **52.81%** | **43.71%** | **1.19** | **1.78** | **54.10%** | **77.92%** | **1.25** | **0.06** | **186.52** | **6.45** | **98.03** | **0.70** |
| ABCA4 | 873 | 7926 | 9 | 46 | 93 | 2.19 | 1.01 | 9.25% | 13.74% | 80.29% | 19.81 | 1.45 | 11.13% | 61.34% | 18.04% | 0.05 | 0.33 | 66.40% | 48.28% | 33.46% | 0.57 | 1.58 | 59.62% | 81.76% | 1.33 | 0.08 | 184.81 | 4.81 | 61.64 | 0.71 |
| ABCC6 | 17 | 268 | 26 | 62 | 90 | 1.53 | 1.03 | 16.85% | 17.58% | 69.03% | 21.44 | 2.08 | 14.31% | 32.63% | 54.85% | 0.23 | 1.20 | 62.52% | 23.36% | 8.21% | 0.03 | 0.15 | 43.31% | 38.00% | 1.37 | 0.08 | 196.68 | 6.17 | 80.72 | 0.73 |
| ABHD12 | 4 | 44 | 26 | 33.5 | 59 | 2.50 | 1.00 | 11.17% | 3.83% | 77.27% | 15.90 | 2.48 | 11.45% | 49.64% | 27.27% | 0.02 | 0.27 | 35.63% | 66.67% | 2.27% | 4.01E-05 | 0.00 | 47.94% | 100.00% | 1.19 | 0.06 | 179.19 | 5.24 | 36.36 | 0.68 |
| ADAMTSL4 | 1 | 12 | 33 | 33 | 33 | 2.27 | 1.00 | 11.05% | 0.00% | 25.00% | 0.02 | 0.33 | 9.49% | 0.00% | 0.00% | 0.00 | 0.00 | | | 16.67% | 0.03 | 0.17 | 42.55% | 100.00% | 1.35 | 0.07 | 213.39 | 4.03 | 53.32 | 0.69 |
| ADGRV1 | 12 | 102 | 26 | 51.5 | 77 | 2.54 | 1.00 | 17.39% | 1.46% | 63.73% | 4.88 | 1.19 | 17.73% | 46.66% | 8.82% | 0.08 | 0.12 | 64.25% | 86.49% | 86.27% | 1.09 | 3.35 | 52.19% | 99.03% | 1.13 | 0.02 | 182.36 | 8.66 | 140.31 | 0.67 |
| AGBL5 | 1 | 10 | 73 | 73 | 73 | 0.54 | 1.60 | 9.81% | 0.00% | 100.00% | 9.49 | 4.80 | 2.21% | 5.33% | 0.00% | 0.00 | 0.00 | | | 0.00% | 0.00 | 0.00 | | | 1.31 | 0.04 | 206.99 | 8.55 | 91.48 | 0.72 |
| AHI1 | 6 | 49 | 17 | 52 | 65 | 2.85 | 1.00 | 10.99% | 5.88% | 65.31% | 12.24 | 0.86 | 12.82% | 53.64% | 10.20% | 0.01 | 0.06 | 42.02% | 60.00% | 42.86% | 0.50 | 0.96 | 54.65% | 85.91% | 1.14 | 0.03 | 180.38 | 5.46 | 58.98 | 0.70 |
| AIPL1 | 2 | 25 | 9 | 15 | 56 | 2.48 | 1.00 | 9.60% | 0.06% | 8.00% | 0.11 | 0.40 | 2.85% | 2.66% | 32.00% | 0.03 | 0.24 | 47.47% | 0.00% | 80.00% | 8.66 | 2.12 | 40.42% | 41.64% | 1.12 | 0.04 | 229.54 | 5.70 | 40.85 | 0.71 |
| ALMS1 | 1 | 11 | 15 | 28 | 48 | 2.36 | 1.00 | 13.89% | 0.00% | 0.00% | 0.00 | 0.00 | | | 0.00% | 0.00 | 0.00 | | | 0.00% | 0.00 | 0.00 | | | 1.10 | 0.04 | 186.00 | 5.01 | 67.32 | 0.70 |
| AMACR | 2 | 12 | 61 | 62.5 | 64 | 1.69 | 1.00 | 18.99% | 0.00% | 41.67% | 0.10 | 0.42 | 13.05% | 77.46% | 66.67% | 0.10 | 0.67 | 56.90% | 3.57% | 25.00% | 0.09 | 0.33 | 43.51% | 75.00% | 1.15 | 0.04 | 191.50 | 7.62 | 110.12 | 0.65 |
| ARHGEF18 | 3 | 39 | 44 | 45 | 60 | 2.34 | 1.00 | 15.63% | 1.72% | 46.15% | 2.88 | 1.13 | 11.33% | 6.43% | 33.33% | 0.12 | 0.69 | 41.63% | 58.53% | 56.41% | 0.99 | 2.08 | 48.92% | 48.09% | 1.22 | 0.05 | 201.54 | 8.35 | 48.53 | 0.64 |
| ARL3 | 1 | 10 | 29 | 29 | 29 | 3.00 | 1.00 | 7.28% | 0.00% | 40.00% | 0.00 | 0.20 | 7.68% | 0.00% | 0.00% | 0.00 | 0.00 | | | 0.00% | 0.00 | 0.00 | | | 1.45 | 0.12 | 198.62 | 4.48 | 27.12 | 0.74 |
| ARL6 | 2 | 14 | 43 | 43.5 | 47 | 1.93 | 1.00 | 21.11% | 22.91% | 92.86% | 35.47 | 1.43 | 16.59% | 46.02% | 50.00% | 0.23 | 0.57 | 43.28% | 85.71% | 0.00% | 0.00 | 0.00 | | | 0.89 | 0.00 | 123.00 | 25.43 | 1151.60 | 0.55 |
| ATF6 | 2 | 15 | 30 | 31.5 | 33 | 2.00 | 1.00 | 17.23% | 0.55% | 53.33% | 2.88 | 0.53 | 13.46% | 81.15% | 46.67% | 0.03 | 0.53 | 55.29% | 38.45% | 13.33% | 0.02 | 0.13 | 29.13% | 100.00% | 1.28 | 0.06 | 229.27 | 4.11 | 47.41 | 0.75 |
| ATXN7 | 1 | 9 | 77 | 77 | 77 | 1.44 | 1.00 | 18.74% | 0.00% | 22.22% | 0.14 | 0.78 | 0.50% | 100.00% | 55.56% | 0.05 | 1.22 | 64.87% | 0.00% | 0.00% | 0.00 | 0.00 | | | 1.36 | 0.08 | 243.18 | 10.45 | 149.14 | 0.73 |
| BBS1 | 31 | 286 | 16 | 38 | 78 | 2.59 | 1.02 | 10.41% | 7.52% | 67.48% | 8.35 | 1.29 | 11.91% | 56.94% | 25.87% | 0.15 | 0.38 | 47.26% | 86.85% | 32.52% | 0.84 | 1.03 | 56.50% | 72.71% | 1.18 | 0.05 | 186.86 | 9.66 | 214.92 | 0.70 |
| BBS10 | 4 | 35 | 17 | 36 | 39 | 2.47 | 1.09 | 12.38% | 30.85% | 91.43% | 8.64 | 1.37 | 11.18% | 41.65% | 0.00% | 0.00 | 0.00 | | | 0.00% | 0.00 | 0.00 | | | 0.96 | 0.01 | 172.29 | 13.56 | 349.29 | 0.68 |
| BBS12 | 2 | 22 | 21 | 35 | 60 | 2.19 | 1.05 | 13.20% | 16.11% | 95.45% | 22.14 | 2.45 | 7.56% | 44.37% | 27.27% | 0.05 | 0.36 | 57.05% | 57.80% | 45.45% | 0.34 | 0.91 | 42.80% | 55.42% | 1.22 | 0.04 | 193.66 | 8.94 | 93.01 | 0.74 |
| BBS2 | 2 | 14 | 36 | 40 | 44 | 2.96 | 1.00 | 7.20% | 19.31% | 100.00% | 4.91 | 1.50 | 14.15% | 76.73% | 0.00% | 0.00 | 0.00 | | | 57.14% | 1.85 | 1.43 | 63.87% | 89.38% | 1.29 | 0.04 | 134.42 | 6.12 | 201.98 | 0.74 |
| BBS5 | 2 | 19 | 32 | 53 | 74 | 3.50 | 1.00 | 12.69% | 3.59% | 5.26% | 0.00 | 0.05 | 23.42% | 0.00% | 31.58% | 0.08 | 0.53 | 46.02% | 82.37% | 84.21% | 0.93 | 3.95 | 47.36% | 80.33% | 1.38 | 0.08 | 206.60 | 11.55 | 199.06 | 0.76 |
| BEST1 | 133 | 1461 | 8 | 46 | 86 | 1.84 | 1.01 | 11.59% | 4.14% | 40.45% | 2.58 | 0.90 | 11.50% | 59.70% | 57.49% | 0.73 | 1.09 | 61.18% | 59.81% | 57.49% | 1.28 | 1.83 | 56.31% | 77.22% | 1.38 | 0.10 | 203.18 | 4.46 | 49.43 | 0.71 |
| C1QTNF5 | 10 | 73 | 50 | 73.5 | 87 | 1.85 | 1.00 | 13.85% | 3.18% | 78.08% | 27.18 | 1.93 | 15.97% | 23.12% | 8.22% | 0.01 | 0.07 | 59.98% | 50.00% | 1.37% | 0.02 | 0.07 | 51.84% | 100.00% | 1.35 | 0.08 | 199.82 | 4.78 | 43.26 | 0.73 |
| C21ORF2 | 5 | 44 | 27 | 37.5 | 57 | 2.56 | 1.00 | 11.52% | 0.37% | 20.45% | 0.76 | 0.25 | 32.97% | 17.62% | 0.00% | 0.00 | 0.00 | | | 52.27% | 1.00 | 1.23 | 56.98% | 45.38% | 1.28 | 0.07 | 191.13 | 4.31 | 52.74 | 0.73 |
| C2ORF71 | 10 | 85 | 52 | 52 | 52 | 2.38 | 1.01 | 13.39% | 6.76% | 96.47% | 19.08 | 1.75 | 12.65% | 65.48% | 7.06% | 0.00 | 0.07 | 55.40% | 83.33% | 23.53% | 0.63 | 0.86 | 62.86% | 62.54% | 1.05 | 0.03 | 177.08 | 13.59 | 252.66 | 0.63 |
| CABP4 | 4 | 29 | 9 | 39 | 48 | 1.33 | 1.03 | 14.68% | 3.45% | 20.69% | 0.01 | 0.79 | 0.03% | 33.33% | 17.24% | 0.00 | 0.21 | 53.22% | 0.00% | 0.00% | 0.00 | 0.00 | | | 1.11 | 0.03 | 224.24 | 2.22 | 20.98 | 0.71 |
| CACNA1F | 30 | 265 | 4 | 18 | 79 | 2.04 | 1.01 | 14.76% | 0.01% | 5.66% | 0.91 | 0.20 | 7.13% | 42.39% | 12.83% | 0.01 | 0.15 | 59.85% | 2.47% | 3.77% | 0.01 | 0.06 | 53.57% | 79.82% | 1.21 | 0.05 | 218.03 | 9.24 | 83.77 | 0.73 |
| CACNA2D4 | 1 | 16 | 40 | 40 | 40 | 1.06 | 1.00 | 17.29% | 0.00% | 100.00% | 0.09 | 4.06 | 0.11% | 80.36% | 0.00% | 0.00 | 0.00 | | | 0.00% | 0.00 | 0.00 | | | 1.49 | 0.10 | 210.24 | 3.97 | 30.23 | 0.69 |
| CDH23 | 17 | 260 | 15 | 40 | 74 | 2.05 | 1.01 | 14.31% | 6.45% | 16.15% | 0.71 | 0.47 | 10.44% | 41.97% | 19.62% | 0.23 | 0.50 | 57.64% | 34.17% | 77.31% | 2.46 | 2.73 | 47.34% | 67.30% | 1.05 | 0.02 | 198.91 | 7.97 | 177.84 | 0.68 |
| CDH3 | 3 | 21 | 32 | 38 | 63 | 1.65 | 1.19 | 15.05% | 0.00% | 100.00% | 25.42 | 2.24 | 11.58% | 48.30% | 42.86% | 0.82 | 1.43 | 45.35% | 0.00% | 28.57% | 0.11 | 1.19 | 43.43% | 0.00% | 1.27 | 0.05 | 219.90 | 3.19 | 17.80 | 0.63 |
| CDHR1 | 18 | 144 | 26 | 41 | 82 | 2.48 | 1.04 | 12.55% | 5.10% | 80.56% | 7.39 | 1.71 | 11.82% | 66.14% | 22.92% | 0.18 | 0.26 | 49.52% | 75.30% | 36.81% | 1.11 | 1.25 | 53.78% | 70.56% | 1.22 | 0.04 | 180.64 | 7.80 | 77.75 | 0.68 |
| CEP290 | 16 | 166 | 6 | 33 | 57 | 2.22 | 0.99 | 14.25% | 3.10% | 19.88% | 0.90 | 0.69 | 6.43% | 48.94% | 13.25% | 0.10 | 0.25 | 62.19% | 13.64% | 70.48% | 1.40 | 2.10 | 54.86% | 80.08% | 1.16 | 0.04 | 186.46 | 10.90 | 281.31 | 0.71 |
| CEP78 | 1 | 5 | 72 | 72 | 72 | 1.47 | 1.00 | 20.86% | 12.22% | 100.00% | 37.38 | 3.80 | 22.08% | 9.46% | 100.00% | 0.56 | 1.80 | 53.42% | 18.37% | 100.00% | 3.94 | 2.00 | 51.21% | 87.14% | 1.01 | 0.01 | 213.43 | 3.91 | 40.37 | 0.77 |
| CERKL | 22 | 249 | 20 | 40 | 80 | 2.63 | 1.00 | 12.26% | 9.15% | 82.33% | 10.33 | 1.43 | 12.08% | 51.71% | 17.67% | 0.02 | 0.21 | 46.03% | 88.64% | 29.72% | 1.04 | 0.95 | 58.07% | 65.30% | 1.16 | 0.04 | 179.55 | 7.20 | 95.84 | 0.67 |
| CHM | 109 | 1731 | 11 | 42 | 86 | 1.45 | 1.06 | 20.61% | 2.84% | 82.61% | 51.77 | 3.28 | 19.89% | 20.95% | 48.76% | 0.26 | 0.83 | 54.55% | 37.86% | 5.55% | 0.03 | 0.08 | 43.19% | 32.21% | 1.30 | 0.06 | 190.34 | 6.14 | 104.36 | 0.69 |
| CLCC1 | 1 | 12 | 25 | 25 | 25 | 2.37 | 1.00 | 22.89% | 0.00% | 25.00% | 0.01 | 0.33 | 12.78% | 0.00% | 0.00% | 0.00 | 0.00 | | | 33.33% | 0.54 | 0.33 | 54.51% | 100.00% | 0.79 | 0.00 | 137.72 | 2.37 | 1.88 | 0.29 |
| CLN3 | 10 | 147 | 13 | 47 | 62 | 2.23 | 1.03 | 16.06% | 8.51% | 34.01% | 1.75 | 1.17 | 13.96% | 37.43% | 2.72% | 0.00 | 0.06 | 57.99% | 25.00% | 47.62% | 0.52 | 1.97 | 49.10% | 92.66% | 1.29 | 0.06 | 217.27 | 7.86 | 143.39 | 0.73 |
| CLRN1 | 10 | 80 | 22 | 47.5 | 75 | 1.40 | 1.04 | 19.90% | 4.06% | 60.00% | 9.38 | 1.71 | 15.71% | 23.13% | 23.75% | 0.06 | 0.41 | 53.97% | 63.16% | 51.25% | 0.97 | 1.44 | 52.49% | 94.12% | 1.02 | 0.01 | 143.88 | 15.66 | 221.72 | 0.67 |
| CNGA1 | 8 | 54 | 34 | 53 | 90 | 2.39 | 1.00 | 18.34% | 2.16% | 16.67% | 2.91 | 0.20 | 20.70% | 75.96% | 3.70% | 0.22 | 0.15 | 59.32% | 21.66% | 22.22% | 1.32 | 0.89 | 60.53% | 65.29% | 1.30 | 0.07 | 206.51 | 7.38 | 89.46 | 0.70 |
| CNGA3 | 36 | 289 | 4 | 23 | 67 | 1.74 | 1.01 | 13.81% | 14.49% | 31.14% | 1.48 | 0.96 | 7.06% | 56.25% | 7.96% | 0.05 | 0.17 | 53.96% | 31.76% | 35.64% | 0.32 | 0.75 | 50.03% | 74.89% | 1.24 | 0.05 | 209.19 | 5.17 | 70.94 | 0.70 |
| CNGB1 | 28 | 198 | 26 | 58 | 88 | 1.59 | 1.03 | 13.07% | 1.74% | 79.80% | 19.98 | 2.03 | 10.08% | 25.60% | 46.46% | 0.57 | 0.95 | 54.04% | 50.81% | 70.71% | 3.63 | 2.64 | 54.52% | 64.45% | 1.17 | 0.03 | 160.01 | 7.17 | 90.08 | 0.68 |
| CNGB3 | 39 | 288 | 5 | 29.5 | 80 | 1.91 | 0.99 | 14.86% | 12.72% | 12.50% | 0.16 | 0.38 | 6.73% | 73.57% | 10.42% | 0.01 | 0.12 | 52.72% | 63.89% | 32.64% | 0.20 | 0.63 | 54.64% | 92.23% | 1.24 | 0.06 | 205.53 | 6.42 | 67.09 | 0.70 |
| COL11A1 | 1 | 8 | 30 | 42 | 56 | 2.64 | 1.00 | 6.77% | 2.55% | 100.00% | 0.73 | 2.75 | 6.21% | 61.71% | 37.50% | 0.01 | 0.38 | 34.48% | 33.33% | 12.50% | 0.04 | 0.38 | 40.40% | 100.00% | 1.13 | 0.04 | 178.64 | 3.60 | 29.13 | 0.58 |
| COL18A1 | 2 | 2 | 9 | 27 | 42 | 0.26 | 1.00 | 13.17% | 0.00% | 50.00% | 34.50 | 3.00 | 19.81% | 14.65% | 50.00% | 0.25 | 0.50 | 55.09% | 1.23% | 0.00% | 0.00 | 0.00 | | | 1.01 | 0.02 | 205.69 | 2.51 | 5.27 | 0.86 |
| COL2A1 | 1 | 8 | 61 | 66 | 71 | 1.71 | 1.38 | 19.62% | 13.38% | 100.00% | 27.44 | 2.63 | 8.53% | 6.64% | 75.00% | 0.56 | 0.88 | 63.15% | 0.00% | 25.00% | 0.07 | 0.25 | 46.61% | 27.85% | 1.15 | 0.03 | 188.28 | 3.09 | 21.79 | 0.69 |
| COL4A1 | 1 | 7 | 48 | 48 | 48 | 1.93 | 1.00 | 16.31% | 0.00% | 0.00% | 0.00 | 0.00 | | | 0.00% | 0.00 | 0.00 | | | 0.00% | 0.00 | 0.00 | | | 1.50 | 0.14 | 188.66 | 3.98 | 37.88 | 0.70 |
| COL4A5 | 1 | 10 | 40 | 40 | 40 | 1.61 | 1.00 | 12.84% | 0.00% | 0.00% | 0.00 | 0.00 | | | 0.00% | 0.00 | 0.00 | | | 100.00% | 11.30 | 4.10 | 57.72% | 80.63% | 1.20 | 0.03 | 194.98 | 8.70 | 85.90 | 0.81 |
| CRB1 | 47 | 455 | 8 | 38 | 80 | 1.97 | 1.02 | 14.70% | 3.63% | 55.16% | 8.30 | 1.22 | 15.60% | 40.44% | 43.96% | 0.41 | 0.89 | 46.39% | 54.00% | 43.74% | 1.73 | 1.35 | 48.91% | 59.21% | 1.11 | 0.04 | 178.82 | 7.20 | 83.21 | 0.69 |
| CRX | 26 | 236 | 20 | 61 | 85 | 2.11 | 1.00 | 15.62% | 5.73% | 70.34% | 5.51 | 1.39 | 15.62% | 62.69% | 16.95% | 0.05 | 0.26 | 50.74% | 29.92% | 59.32% | 2.23 | 4.22 | 52.94% | 65.95% | 1.33 | 0.07 | 192.40 | 9.58 | 119.49 | 0.72 |
| CTNNA1 | 4 | 10 | 33 | 39 | 65 | 1.51 | 1.30 | 18.54% | 0.00% | 40.00% | 1.50 | 2.20 | 1.60% | 13.10% | 60.00% | 0.23 | 1.20 | 56.33% | 99.44% | 50.00% | 0.04 | 0.80 | 56.60% | 80.51% | 1.42 | 0.09 | 192.93 | 4.01 | 26.94 | 0.71 |
| CWC27 | 1 | 1 | 19 | 19 | 19 | 0.30 | 1.00 | 21.28% | 0.00% | 0.00% | 0.00 | 0.00 | | | 0.00% | 0.00 | 0.00 | | | 0.00% | 0.00 | 0.00 | | | 0.56 | 0.00 | 80.80 | 3.13 | 1.54 | 0.41 |
| CYP4V2 | 17 | 106 | 21 | 47 | 86 | 1.18 | 1.06 | 22.53% | 3.07% | 86.79% | 35.05 | 3.20 | 16.02% | 33.68% | 15.09% | 0.02 | 0.20 | 49.14% | 61.88% | 3.77% | 0.01 | 0.04 | 32.83% | 50.00% | 1.00 | 0.02 | 144.85 | 4.90 | 28.37 | 0.63 |
| DRAM2 | 8 | 70 | 43 | 57 | 74 | 2.46 | 1.00 | 14.67% | 9.78% | 90.00% | 5.97 | 1.50 | 17.68% | 89.28% | 2.86% | 0.00 | 0.03 | 29.53% | 100.00% | 34.29% | 0.29 | 0.96 | 59.39% | 73.82% | 1.37 | 0.08 | 190.91 | 5.03 | 65.16 | 0.73 |
| DYNC2H1 | 1 | 3 | 51 | 51 | 51 | 3.05 | 1.00 | 33.60% | 0.00% | 33.33% | 0.30 | 0.33 | 35.71% | 100.00% | 0.00% | 0.00 | 0.00 | | | 66.67% | 0.76 | 1.67 | 61.57% | 100.00% | 1.07 | 0.02 | 155.01 | 7.69 | 157.23 | 0.49 |
| EFEMP1 | 30 | 249 | 26 | 58.5 | 97 | 1.77 | 1.00 | 15.28% | 5.98% | 78.71% | 5.15 | 1.84 | 14.24% | 82.36% | 77.91% | 0.24 | 1.55 | 61.84% | 79.37% | 24.10% | 0.18 | 0.66 | 60.03% | 82.55% | 1.43 | 0.10 | 208.55 | 4.31 | 39.45 | 0.72 |
| EYS | 70 | 628 | 10 | 59 | 88 | 2.36 | 1.02 | 15.16% | 2.88% | 65.92% | 9.55 | 1.64 | 13.27% | 27.28% | 23.41% | 0.23 | 0.40 | 57.64% | 42.90% | 72.45% | 2.08 | 2.76 | 52.59% | 76.78% | 1.16 | 0.03 | 172.31 | 7.39 | 108.29 | 0.69 |
| FAM161A | 8 | 62 | 27 | 50 | 63 | 2.12 | 1.05 | 14.58% | 14.56% | 66.13% | 11.60 | 2.02 | 13.90% | 40.33% | 19.35% | 0.27 | 0.35 | 46.78% | 58.46% | 83.87% | 1.83 | 2.77 | 51.04% | 90.53% | 1.10 | 0.02 | 168.97 | 5.66 | 45.07 | 0.65 |
| FLVCR1 | 4 | 27 | 52 | 53 | 59 | 1.01 | 0.93 | 16.45% | 3.53% | 48.15% | 2.58 | 1.85 | 5.93% | 41.36% | 18.52% | 0.15 | 0.37 | 75.14% | 20.00% | 74.07% | 3.76 | 1.78 | 49.80% | 78.55% | 1.06 | 0.02 | 176.65 | 9.58 | 88.25 | 0.70 |
| FZD4 | 2 | 9 | 9 | 14 | 67 | 2.78 | 1.11 | 9.17% | 16.13% | 66.67% | 2.59 | 1.11 | 9.48% | 58.83% | 44.44% | 0.12 | 0.56 | 78.35% | 0.00% | 33.33% | 0.02 | 0.56 | 71.29% | 0.00% | 1.40 | 0.10 | 211.97 | 4.37 | 35.44 | 0.66 |
| GNAT2 | 2 | 31 | 30 | 32 | 34 | 2.03 | 1.00 | 15.64% | 0.00% | 0.00% | 0.00 | 0.00 | | | 9.68% | 0.01 | 0.10 | 52.77% | 0.00% | 0.00% | 0.00 | 0.00 | | | 1.35 | 0.08 | 230.33 | 4.50 | 55.07 | 0.73 |

| | | | | | | Disc | | | | Hypo-AF | | | | | Hyper-AF | | | | | Ring | | | | | Vessels | | | | | |
|---|---|---|---|---|---|---|---|---|---|---|---|---|---|---|---|---|---|---|---|---|---|---|---|---|---|---|---|---|---|---|
| gene | patient count | image count | Patient Ages | | | area | num clusters | intensity | % <6mm | incidence | area | num clusters | intensity | % <6mm | incidence | area | num clusters | intensity | % <6mm | incidence | area | num clusters | intensity | % <6mm | Fractal Dimension | Density | Width | Distance tortuosity mean | Squared curvature tortuosity mean | Tortuosity density mean |
| **Total** | **3491** | **32964** | **min** | **median** | **max** | **2.09** | **1.02** | **13.51%** | **6.86%** | **61.15%** | **13.04** | **1.46** | **13.02%** | **48.17%** | **25.11%** | **0.15** | **0.43** | **57.14%** | **52.81%** | **43.71%** | **1.19** | **1.78** | **54.10%** | **77.92%** | **1.25** | **0.06** | **186.52** | **6.45** | **98.03** | **0.70** |
| GPR143 | 6 | 28 | 14 | 35 | 69 | 2.31 | 1.00 | 13.46% | 0.74% | 0.00% | 0.00 | 0.00 | | | 0.00% | 0.00 | 0.00 | | | 0.00% | 0.00 | 0.00 | | | 1.43 | 0.11 | 209.76 | 5.46 | 61.22 | 0.71 |
| GPR179 | 2 | 12 | 13 | 25.5 | 40 | 2.71 | 1.00 | 13.40% | 3.57% | 50.00% | 0.95 | 0.67 | 21.58% | 100.00% | 0.00% | 0.00 | 0.00 | | | 50.00% | 1.97 | 1.33 | 51.76% | 100.00% | 1.30 | 0.06 | 221.14 | 9.26 | 157.48 | 0.75 |
| GRK1 | 1 | 4 | 48 | 48 | 48 | 3.41 | 1.00 | 22.44% | 0.00% | 100.00% | 2.02 | 1.00 | 31.29% | 97.92% | 0.00% | 0.00 | 0.00 | | | 100.00% | 1.88 | 2.25 | 54.96% | 100.00% | 1.28 | 0.04 | 185.11 | 8.14 | 150.90 | 0.76 |
| GRM6 | 5 | 17 | 9 | 27 | 59 | 1.69 | 1.12 | 14.61% | 0.00% | 41.18% | 0.42 | 0.82 | 8.63% | 9.56% | 23.53% | 0.04 | 0.24 | 60.87% | 0.00% | 11.76% | 0.02 | 0.18 | 73.19% | 0.00% | 1.19 | 0.06 | 162.84 | 3.47 | 23.22 | 0.66 |
| GUCA1A | 11 | 79 | 29 | 58 | 73 | 2.25 | 1.01 | 16.55% | 9.69% | 41.77% | 2.57 | 0.80 | 17.77% | 58.47% | 25.32% | 0.02 | 0.24 | 57.05% | 65.00% | 69.62% | 1.14 | 1.89 | 60.80% | 93.77% | 1.21 | 0.07 | 175.35 | 4.35 | 34.07 | 0.61 |
| GUCY2D | 28 | 256 | 13 | 45 | 74 | 2.00 | 1.00 | 14.65% | 6.51% | 48.05% | 5.25 | 0.86 | 17.50% | 71.23% | 17.58% | 0.22 | 0.30 | 61.24% | 35.99% | 60.94% | 1.51 | 2.94 | 53.11% | 74.98% | 1.29 | 0.07 | 197.87 | 5.41 | 64.41 | 0.72 |
| HGSNAT | 11 | 150 | 53 | 69 | 86 | 2.04 | 1.01 | 19.73% | 7.15% | 87.33% | 25.58 | 2.53 | 19.35% | 23.44% | 47.33% | 0.61 | 0.89 | 59.42% | 60.81% | 76.00% | 3.46 | 2.91 | 50.43% | 84.38% | 1.16 | 0.04 | 173.35 | 6.01 | 52.57 | 0.68 |
| HPS6 | 1 | 2 | 7 | 10 | 57 | 1.98 | 1.00 | 10.33% | 0.00% | 0.00% | 0.00 | 0.00 | | | 50.00% | 0.01 | 0.50 | 60.38% | 0.00% | 100.00% | 0.66 | 1.50 | 67.26% | 91.78% | 1.43 | 0.12 | 224.18 | 5.33 | 53.82 | 0.70 |
| IFT140 | 8 | 74 | 0 | 46.5 | 95 | 2.45 | 1.01 | 13.92% | 2.45% | 72.97% | 26.72 | 1.93 | 14.83% | 41.25% | 28.38% | 0.04 | 0.34 | 51.37% | 54.63% | 27.03% | 0.57 | 0.77 | 47.30% | 63.40% | 1.05 | 0.03 | 178.13 | 8.60 | 175.65 | 0.64 |
| IMPDH1 | 6 | 102 | 18 | 60 | 85 | 1.03 | 1.15 | 20.24% | 1.41% | 16.67% | 0.74 | 0.49 | 7.60% | 6.21% | 70.59% | 0.69 | 1.00 | 78.22% | 1.37% | 74.51% | 5.84 | 1.47 | 43.96% | 42.58% | 1.10 | 0.03 | 186.45 | 11.70 | 355.53 | 0.72 |
| IMPG1 | 2 | 10 | 45 | 66 | 88 | 2.57 | 1.00 | 13.09% | 10.11% | 70.00% | 0.63 | 0.60 | 10.29% | 100.00% | 30.00% | 0.16 | 0.30 | 62.18% | 66.67% | 70.00% | 0.25 | 1.60 | 61.95% | 53.47% | 1.41 | 0.10 | 206.75 | 9.02 | 147.19 | 0.72 |
| IMPG2 | 12 | 101 | 26 | 40 | 75 | 2.61 | 1.02 | 14.77% | 3.69% | 58.42% | 4.78 | 1.06 | 13.69% | 30.79% | 9.90% | 0.05 | 0.21 | 57.06% | 57.45% | 27.72% | 0.26 | 0.71 | 44.02% | 68.09% | 1.13 | 0.03 | 197.91 | 7.92 | 135.19 | 0.68 |
| INPP5E | 4 | 27 | 28 | 33.5 | 47 | 1.88 | 1.04 | 9.15% | 3.72% | 37.04% | 0.95 | 1.85 | 1.51% | 35.60% | 7.41% | 0.00 | 0.07 | 42.62% | 100.00% | 92.59% | 3.00 | 4.00 | 58.85% | 80.90% | 1.38 | 0.07 | 197.45 | 4.24 | 56.23 | 0.68 |
| IQCB1 | 8 | 48 | 13 | 45 | 80 | 2.33 | 1.00 | 10.80% | 18.66% | 18.75% | 1.36 | 0.27 | 14.41% | 34.34% | 14.58% | 0.10 | 0.19 | 41.52% | 51.81% | 85.42% | 3.57 | 3.42 | 38.49% | 71.76% | 1.19 | 0.04 | 214.53 | 4.42 | 35.61 | 0.71 |
| JAG1 | 1 | 10 | 36 | 36 | 36 | 2.85 | 1.00 | 12.21% | 52.78% | 0.00% | 0.00 | 0.00 | | | 0.00% | 0.00 | 0.00 | | | 0.00% | 0.00 | 0.00 | | | 1.41 | 0.09 | 195.27 | 6.02 | 179.10 | 0.77 |
| KCNJ13 | 1 | 1 | 44 | 44 | 44 | 0.04 | 1.00 | 6.74% | 0.00% | 100.00% | 24.14 | 1.00 | 7.38% | 51.12% | 0.00% | 0.00 | 0.00 | | | 0.00% | 0.00 | 0.00 | | | 0.78 | 0.00 | 132.76 | 11.98 | 34.13 | 0.51 |
| KCNV2 | 24 | 159 | 9 | 35 | 77 | 2.06 | 1.01 | 12.71% | 5.01% | 35.85% | 1.77 | 0.75 | 13.62% | 84.52% | 9.43% | 0.01 | 0.08 | 53.98% | 67.72% | 52.20% | 0.92 | 2.87 | 51.85% | 82.53% | 1.27 | 0.07 | 199.15 | 6.01 | 79.68 | 0.73 |
| KIF11 | 1 | 1 | 10 | 14 | 18 | 1.55 | 1.00 | 16.73% | 0.00% | 0.00% | 0.00 | 0.00 | | | 0.00% | 0.00 | 0.00 | | | 0.00% | 0.00 | 0.00 | | | 0.83 | 0.00 | 158.06 | 1.13 | 1.25 | 0.67 |
| KIZ | 1 | 2 | 22 | 22 | 22 | 2.77 | 1.00 | 8.86% | 0.00% | 0.00% | 0.00 | 0.00 | | | 0.00% | 0.00 | 0.00 | | | 100.00% | 2.97 | 9.50 | 46.16% | 100.00% | 1.41 | 0.10 | 108.94 | 8.33 | 73.84 | 0.77 |
| KLHL7 | 8 | 75 | 31 | 44 | 80 | 2.40 | 0.99 | 9.63% | 0.22% | 70.67% | 13.00 | 1.92 | 13.58% | 30.31% | 24.00% | 0.29 | 0.32 | 55.77% | 93.11% | 84.00% | 3.32 | 5.05 | 49.44% | 89.12% | 1.08 | 0.02 | 155.27 | 4.88 | 46.52 | 0.68 |
| LAMA1 | 3 | 64 | 39 | 42 | 44 | 1.82 | 1.03 | 15.16% | 11.46% | 73.44% | 14.25 | 1.59 | 11.84% | 3.88% | 62.50% | 0.68 | 1.02 | 51.90% | 29.27% | 68.75% | 1.70 | 3.20 | 49.59% | 34.35% | 1.28 | 0.05 | 230.37 | 2.95 | 26.37 | 0.79 |
| LCA3 | 1 | 2 | 29 | 29 | 29 | 0.21 | 1.00 | 8.14% | 0.00% | 0.00% | 0.00 | 0.00 | | | 0.00% | 0.00 | 0.00 | | | 100.00% | 1.05 | 4.00 | 23.52% | 90.24% | 0.89 | 0.00 | 173.64 | 2.87 | 41.26 | 0.69 |
| LCA5 | 3 | 22 | 11 | 31 | 41 | 1.68 | 0.95 | 12.37% | 1.64% | 77.27% | 21.91 | 0.95 | 12.25% | 36.27% | 0.00% | 0.00 | 0.00 | | | 0.00% | 0.00 | 0.00 | | | 0.91 | 0.01 | 163.08 | 2.27 | 11.30 | 0.61 |
| LHON | 3 | 9 | 10 | 56 | 77 | 1.93 | 1.00 | 16.85% | 11.09% | 22.22% | 0.26 | 0.44 | 13.18% | 0.00% | 22.22% | 0.01 | 0.22 | 30.78% | 0.00% | 22.22% | 0.05 | 0.44 | 30.19% | 0.00% | 1.29 | 0.07 | 200.32 | 11.21 | 156.87 | 0.75 |
| LRAT | 1 | 6 | 45 | 56.5 | 68 | 1.27 | 1.00 | 47.65% | 0.00% | 0.00% | 0.00 | 0.00 | | | 83.33% | 0.18 | 1.00 | 65.92% | 38.33% | 50.00% | 0.10 | 0.50 | 67.11% | 67.84% | 0.93 | 0.01 | 198.61 | 64.14 | 3810.75 | 0.84 |
| MAK | 1 | 6 | 60 | 60 | 60 | 1.97 | 1.00 | 22.73% | 0.00% | 100.00% | 2.96 | 3.17 | 15.83% | 19.65% | 0.00% | 0.00 | 0.00 | | | 50.00% | 0.48 | 2.17 | 47.06% | 97.53% | 1.12 | 0.01 | 163.64 | 8.17 | 284.51 | 0.70 |
| MERTK | 16 | 199 | 17 | 29 | 62 | 2.32 | 1.01 | 16.10% | 2.09% | 50.25% | 4.65 | 0.78 | 13.72% | 67.63% | 40.70% | 0.09 | 0.58 | 50.57% | 74.20% | 49.25% | 0.26 | 1.11 | 52.78% | 86.12% | 1.12 | 0.03 | 203.52 | 7.47 | 143.86 | 0.70 |
| MFRP | 5 | 95 | 11 | 36.5 | 57 | 1.52 | 1.05 | 9.39% | 16.23% | 63.16% | 4.52 | 1.24 | 5.81% | 12.76% | 49.47% | 0.14 | 0.84 | 66.66% | 11.94% | 10.53% | 0.11 | 0.38 | 35.89% | 90.00% | 1.45 | 0.12 | 219.12 | 4.58 | 59.51 | 0.72 |
| MFSD8 | 12 | 132 | 35 | 52 | 86 | 2.50 | 1.02 | 16.29% | 11.16% | 90.91% | 9.35 | 1.48 | 16.31% | 77.20% | 4.55% | 0.00 | 0.06 | 53.64% | 83.33% | 29.55% | 0.71 | 1.06 | 53.56% | 79.47% | 1.16 | 0.05 | 180.60 | 4.76 | 37.14 | 0.68 |
| MKKS | 2 | 41 | 21 | 23 | 25 | 2.79 | 1.07 | 10.91% | 23.12% | 73.17% | 0.82 | 1.10 | 13.35% | 61.34% | 58.54% | 0.13 | 0.59 | 41.18% | 57.57% | 0.00% | 0.00 | 0.00 | | | 0.99 | 0.01 | 199.05 | 17.37 | 849.58 | 0.71 |
| MT-ATP6 | 2 | 30 | 39 | 49 | 59 | 1.87 | 1.00 | 8.18% | 0.02% | 100.00% | 14.49 | 2.27 | 11.39% | 38.65% | 76.67% | 0.64 | 1.57 | 38.90% | 71.42% | 50.00% | 0.25 | 0.57 | 37.80% | 83.24% | 0.90 | 0.01 | 142.39 | 15.17 | 584.20 | 0.73 |
| MT-ND1, MT-N | 2 | 11 | 25 | 51 | 82 | 1.94 | 1.00 | 22.93% | 0.00% | 0.00% | 0.00 | 0.00 | | | 0.00% | 0.00 | 0.00 | | | 0.00% | 0.00 | 0.00 | | | 1.40 | 0.10 | 204.03 | 6.87 | 77.56 | 0.74 |
| MT-TL1 | 18 | 121 | 34 | 61 | 82 | 2.45 | 1.00 | 15.52% | 9.29% | 99.17% | 27.19 | 1.83 | 17.96% | 60.58% | 8.26% | 0.01 | 0.09 | 72.61% | 53.87% | 2.48% | 0.03 | 0.05 | 57.33% | 41.27% | 1.38 | 0.08 | 200.53 | 4.72 | 56.76 | 0.73 |
| MYO7A | 49 | 489 | 12 | 45 | 85 | 2.07 | 0.99 | 15.77% | 2.33% | 42.33% | 2.37 | 0.95 | 18.00% | 32.93% | 28.02% | 0.41 | 0.97 | 53.69% | 21.82% | 73.62% | 3.24 | 3.26 | 47.00% | 57.83% | 1.09 | 0.03 | 182.09 | 11.00 | 296.77 | 0.70 |
| NDP | 1 | 4 | 37 | 37.5 | 38 | 2.60 | 1.00 | 6.09% | 25.00% | 50.00% | 10.17 | 2.50 | 6.10% | 0.00% | 50.00% | 0.05 | 0.50 | 68.53% | 50.00% | 0.00% | 0.00 | 0.00 | | | 1.31 | 0.09 | 193.26 | 4.32 | 60.09 | 0.72 |
| NEK1 | 1 | 2 | 29 | 29 | 29 | 2.26 | 1.00 | 9.98% | 0.00% | 0.00% | 0.00 | 0.00 | | | 50.00% | 0.01 | 0.50 | 50.11% | 0.00% | 0.00% | 0.00 | 0.00 | | | 1.44 | 0.11 | 213.59 | 2.76 | 17.96 | 0.72 |
| NHS | 1 | 5 | 49 | 49 | 49 | 2.52 | 1.00 | 13.94% | 2.33% | 0.00% | 0.00 | 0.00 | | | 0.00% | 0.00 | 0.00 | | | 0.00% | 0.00 | 0.00 | | | 1.39 | 0.09 | 214.62 | 5.06 | 34.29 | 0.75 |
| NMNAT1 | 1 | 6 | 12 | 21 | 66 | 2.15 | 1.00 | 22.95% | 0.00% | 100.00% | 6.93 | 2.33 | 25.50% | 11.76% | 100.00% | 2.91 | 2.67 | 70.54% | 0.00% | 100.00% | 2.20 | 2.67 | 60.04% | 11.90% | 1.16 | 0.04 | 208.21 | 9.04 | 31.13 | 0.83 |
| NPHP4 | 1 | 9 | 59 | 59 | 59 | 2.20 | 1.00 | 19.19% | 0.00% | 0.00% | 0.00 | 0.00 | | | 55.56% | 0.04 | 0.44 | 64.42% | 0.00% | 100.00% | 6.05 | 1.67 | 57.05% | 60.07% | 1.24 | 0.04 | 225.33 | 7.56 | 204.69 | 0.70 |
| NR2E3 | 27 | 384 | 9 | 40 | 70 | 1.94 | 1.02 | 10.88% | 7.39% | 31.51% | 1.42 | 0.76 | 10.23% | 12.89% | 21.35% | 0.37 | 0.50 | 56.46% | 17.86% | 39.58% | 1.72 | 1.40 | 52.18% | 24.68% | 1.32 | 0.08 | 202.83 | 5.85 | 77.48 | 0.72 |
| NR2F1 | 1 | 5 | 22 | 29 | 32 | 1.87 | 1.00 | 11.70% | 0.00% | 0.00% | 0.00 | 0.00 | | | 0.00% | 0.00 | 0.00 | | | 0.00% | 0.00 | 0.00 | | | 1.32 | 0.08 | 209.49 | 15.72 | 110.93 | 0.65 |
| NRL | 6 | 26 | 20 | 47 | 80 | 2.01 | 1.04 | 16.78% | 8.27% | 69.23% | 8.00 | 2.19 | 13.37% | 41.91% | 26.92% | 0.12 | 0.31 | 56.64% | 88.18% | 69.23% | 2.80 | 1.85 | 48.05% | 90.56% | 1.16 | 0.03 | 201.74 | 8.27 | 54.69 | 0.68 |
| NYX | 7 | 46 | 5 | 15.5 | 46 | 2.39 | 1.00 | 13.23% | 1.03% | 4.35% | 0.20 | 0.26 | 2.38% | 57.84% | 4.35% | 0.00 | 0.04 | 65.70% | 0.00% | 13.04% | 0.36 | 0.28 | 59.71% | 40.38% | 1.22 | 0.07 | 202.06 | 5.87 | 44.63 | 0.72 |
| OAT | 8 | 143 | 22 | 43.5 | 64 | 1.93 | 1.14 | 11.96% | 1.96% | 74.83% | 47.65 | 2.17 | 9.98% | 12.27% | 61.54% | 0.22 | 1.01 | 51.11% | 20.33% | 18.88% | 0.12 | 0.31 | 35.56% | 12.35% | 1.13 | 0.03 | 191.12 | 7.10 | 85.81 | 0.67 |
| OCA1 | 1 | 1 | 9 | 25.5 | 42 | 0.97 | 1.00 | 10.58% | 0.00% | 0.00% | 0.00 | 0.00 | | | 0.00% | 0.00 | 0.00 | | | 0.00% | 0.00 | 0.00 | | | 0.93 | 0.02 | 189.77 | 2.07 | 30.82 | 0.52 |
| OCA2 | 8 | 79 | 4 | 27 | 83 | 1.64 | 1.01 | 24.24% | 1.33% | 7.59% | 0.99 | 0.16 | 27.76% | 3.03% | 3.80% | 0.00 | 0.04 | 75.84% | 0.00% | 0.00% | 0.00 | 0.00 | | | 1.05 | 0.02 | 202.58 | 6.34 | 42.42 | 0.65 |
| OPA1 | 26 | 127 | 11 | 36 | 79 | 1.89 | 1.00 | 17.48% | 5.24% | 1.57% | 0.01 | 0.02 | 12.34% | 0.00% | 16.54% | 0.01 | 0.30 | 57.81% | 8.61% | 0.79% | 0.00 | 0.01 | 68.94% | 0.00% | 1.40 | 0.10 | 210.36 | 5.59 | 71.17 | 0.73 |
| OPN1LW | 1 | 2 | 31 | 44 | 57 | 2.17 | 1.00 | 10.64% | 0.00% | 0.00% | 0.00 | 0.00 | | | 50.00% | 0.25 | 1.00 | 55.91% | 0.00% | 0.00% | 0.00 | 0.00 | | | 1.46 | 0.12 | 181.40 | 3.60 | 39.21 | 0.74 |
| PAX2 | 1 | 10 | 33 | 39 | 45 | 2.06 | 1.00 | 30.18% | 0.00% | 10.00% | 1.97 | 0.10 | 24.27% | 23.62% | 10.00% | 0.01 | 0.10 | 77.32% | 0.00% | 10.00% | 0.00 | 0.10 | 74.14% | 0.00% | 1.12 | 0.03 | 207.97 | 4.36 | 28.24 | 0.53 |
| PAX6 | 7 | 27 | 11 | 29 | 75 | 1.78 | 1.00 | 15.30% | 0.00% | 0.00% | 0.00 | 0.00 | | | 7.41% | 0.00 | 0.00 | 70.69% | | 0.00% | 0.00 | 0.00 | | | 1.08 | 0.04 | 195.64 | 7.60 | 43.89 | 0.69 |
| PCDH15 | 8 | 103 | 19 | 52 | 81 | 2.17 | 1.04 | 14.76% | 0.00% | 74.76% | 9.68 | 2.16 | 10.91% | 24.24% | 33.01% | 0.15 | 0.47 | 56.83% | 62.04% | 87.38% | 4.50 | 3.33 | 51.23% | 65.06% | 1.12 | 0.02 | 162.39 | 6.92 | 98.70 | 0.67 |
| PDE6A | 13 | 108 | 22 | 36 | 85 | 2.61 | 1.02 | 16.48% | 0.02% | 52.78% | 12.80 | 1.38 | 17.06% | 23.69% | 35.19% | 0.19 | 0.51 | 49.45% | 67.39% | 65.74% | 2.14 | 3.56 | 49.36% | 95.54% | 1.09 | 0.03 | 197.01 | 10.30 | 197.25 | 0.71 |
| PDE6B | 24 | 272 | 10 | 41.5 | 87 | 2.62 | 1.01 | 14.75% | 3.35% | 52.21% | 4.48 | 0.93 | 17.05% | 49.30% | 21.32% | 0.08 | 0.28 | 55.09% | 56.89% | 86.03% | 3.37 | 4.06 | 47.35% | 86.55% | 1.12 | 0.03 | 181.70 | 7.22 | 113.36 | 0.71 |
| PDE6C | 7 | 66 | 9 | 43 | 58 | 2.17 | 1.00 | 14.32% | 6.53% | 40.91% | 1.21 | 0.52 | 11.44% | 79.35% | 4.55% | 0.00 | 0.03 | 47.63% | 100.00% | 45.45% | 1.25 | 2.65 | 45.12% | 68.89% | 1.14 | 0.03 | 209.81 | 10.06 | 141.35 | 0.66 |
| PDE6G | 2 | 17 | 46 | 61.5 | 77 | 1.93 | 1.00 | 14.42% | 0.00% | 94.12% | 24.60 | 2.06 | 8.28% | 2.59% | 5.88% | 0.06 | 0.06 | 42.42% | 100.00% | 58.82% | 0.89 | 0.88 | 52.89% | 100.00% | 0.93 | 0.01 | 146.18 | 8.57 | 97.44 | 0.63 |
| PEX1 | 1 | 1 | 19 | 19 | 19 | 1.12 | 1.00 | 16.41% | 0.00% | 0.00% | 0.00 | 0.00 | | | 0.00% | 0.00 | 0.00 | | | 0.00% | 0.00 | 0.00 | | | 0.96 | 0.01 | 169.76 | 5.39 | 20.10 | 0.79 |
| PHYH | 3 | 23 | 36 | 42 | 43 | 1.69 | 1.04 | 16.25% | 7.65% | 73.91% | 15.09 | 2.00 | 16.61% | 49.14% | 17.39% | 0.13 | 0.22 | 48.62% | 70.92% | 56.52% | 2.32 | 1.57 | 52.79% | 55.29% | 1.02 | 0.02 | 211.91 | 19.76 | 587.68 | 0.70 |
| PMM2 | 1 | 8 | 50 | 50 | 50 | 3.17 | 1.00 | 9.85% | 0.00% | 50.00% | 0.51 | 0.50 | 16.16% | 100.00% | 0.00% | 0.00 | 0.00 | | | 50.00% | 1.49 | 1.50 | 43.02% | 100.00% | 1.32 | 0.07 | 189.42 | 3.90 | 34.53 | 0.72 |
| PNPLA6 | 4 | 44 | 13 | 27 | 64 | 1.09 | 0.95 | 17.85% | 37.45% | 90.91% | 76.83 | 2.05 | 16.65% | 41.66% | 11.36% | 0.03 | 0.14 | 32.67% | 60.43% | 20.45% | 1.52 | 3.11 | 45.99% | 77.18% | 1.11 | 0.03 | 212.07 | 12.77 | 169.49 | 0.70 |

| | | | | | | Disc | | | | Hypo-AF | | | | | Hyper-AF | | | | | Ring | | | | | Vessels | | | | | |
|---|---|---|---|---|---|---|---|---|---|---|---|---|---|---|---|---|---|---|---|---|---|---|---|---|---|---|---|---|---|---|
| gene | patient count | image count | Patient Ages | | | area | num clusters | intensity | % <6mm | incidence | area | num clusters | intensity | % <6mm | incidence | area | num clusters | intensity | % <6mm | incidence | area | num clusters | intensity | % <6mm | Fractal Dimension | Density | Width | Distance tortuosity mean | Squared curvature tortuosity mean | Tortuosity density mean |
| **Total** | **3491** | **32964** | **min** | **median** | **max** | **2.09** | **1.02** | **13.51%** | **6.86%** | **61.15%** | **13.04** | **1.46** | **13.02%** | **48.17%** | **25.11%** | **0.15** | **0.43** | **57.14%** | **52.81%** | **43.71%** | **1.19** | **1.78** | **54.10%** | **77.92%** | **1.25** | **0.06** | **186.52** | **6.45** | **98.03** | **0.70** |
| POC1B | 4 | 39 | 30 | 38 | 59 | 2.01 | 1.00 | 15.65% | 6.44% | 12.82% | 0.80 | 0.13 | 24.00% | 99.92% | 2.56% | 0.00 | 0.03 | 83.77% | 0.00% | 7.69% | 0.17 | 0.41 | 58.99% | 71.49% | 1.33 | 0.08 | 213.24 | 8.42 | 149.54 | 0.73 |
| PROM1 | 52 | 461 | 12 | 46 | 86 | 2.31 | 1.01 | 12.44% | 6.86% | 80.69% | 12.22 | 1.73 | 12.48% | 72.05% | 20.61% | 0.03 | 0.25 | 52.15% | 72.15% | 34.49% | 0.91 | 2.02 | 53.90% | 82.54% | 1.26 | 0.06 | 180.39 | 6.32 | 113.25 | 0.72 |
| PRPF3 | 6 | 68 | 19 | 42 | 87 | 2.19 | 0.99 | 19.71% | 4.63% | 73.53% | 3.76 | 1.07 | 13.84% | 67.07% | 45.59% | 0.61 | 0.50 | 56.20% | 86.15% | 76.47% | 3.17 | 2.24 | 46.35% | 72.56% | 1.03 | 0.02 | 181.57 | 9.51 | 132.07 | 0.72 |
| PRPF31 | 57 | 592 | 12 | 46 | 95 | 2.20 | 1.03 | 15.53% | 1.24% | 48.14% | 6.78 | 1.56 | 11.97% | 41.34% | 18.75% | 0.12 | 0.34 | 45.89% | 68.33% | 64.02% | 2.17 | 2.85 | 52.53% | 86.92% | 1.21 | 0.04 | 178.26 | 7.11 | 146.19 | 0.70 |
| PRPF6 | 4 | 13 | 45 | 58 | 59 | 2.41 | 1.00 | 21.36% | 13.61% | 76.92% | 7.04 | 1.15 | 19.73% | 16.43% | 30.77% | 0.11 | 0.69 | 46.10% | 81.05% | 53.85% | 4.38 | 0.54 | 44.78% | 40.87% | 1.20 | 0.04 | 189.15 | 6.06 | 67.83 | 0.72 |
| PRPF8 | 18 | 172 | 20 | 51 | 83 | 2.33 | 1.02 | 14.57% | 2.46% | 29.65% | 1.73 | 0.65 | 10.35% | 26.14% | 13.95% | 0.02 | 0.15 | 62.66% | 44.31% | 86.05% | 2.78 | 2.72 | 52.30% | 88.57% | 1.12 | 0.03 | 167.83 | 10.67 | 145.07 | 0.69 |
| PRPH2 | 148 | 1218 | 9 | 59 | 91 | 2.05 | 1.01 | 12.34% | 4.47% | 68.72% | 10.28 | 1.89 | 10.10% | 62.00% | 36.37% | 0.07 | 0.58 | 59.10% | 77.08% | 26.27% | 0.69 | 0.85 | 55.77% | 69.43% | 1.36 | 0.08 | 184.62 | 4.76 | 52.09 | 0.71 |
| PRSS56 | 1 | 2 | 30 | 30 | 30 | 1.98 | 1.00 | 22.54% | 11.52% | 0.00% | 0.00 | 0.00 | | | 0.00% | 0.00 | 0.00 | | | 0.00% | 0.00 | 0.00 | | | 1.42 | 0.13 | 262.82 | 2.82 | 18.30 | 0.72 |
| PYGM | 3 | 14 | 50 | 73 | 80 | 2.21 | 1.00 | 17.53% | 0.00% | 21.43% | 3.02 | 0.21 | 10.63% | 82.58% | 71.43% | 0.09 | 1.14 | 62.44% | 81.32% | 0.00% | 0.00 | 0.00 | | | 1.42 | 0.11 | 145.64 | 3.71 | 29.34 | 0.74 |
| RAB28 | 1 | 32 | 12 | 12 | 12 | 2.63 | 1.00 | 11.14% | 0.00% | 0.00% | 0.00 | 0.00 | | | 46.88% | 0.13 | 0.47 | 72.50% | 100.00% | 100.00% | 1.32 | 1.41 | 69.75% | 100.00% | 1.32 | 0.06 | 218.28 | 4.60 | 118.18 | 0.69 |
| RAX2 | 3 | 40 | 45 | 49 | 70 | 2.58 | 1.00 | 20.76% | 19.89% | 90.00% | 6.76 | 1.05 | 22.01% | 52.94% | 7.50% | 0.02 | 0.10 | 62.30% | 34.82% | 32.50% | 0.07 | 0.63 | 48.86% | 77.67% | 1.26 | 0.05 | 208.85 | 10.22 | 118.59 | 0.70 |
| RBP3 | 2 | 17 | 25 | 28.5 | 32 | 2.06 | 1.00 | 15.14% | 1.73% | 70.59% | 7.06 | 1.35 | 13.79% | 34.18% | 52.94% | 0.67 | 1.53 | 45.94% | 22.22% | 11.76% | 0.01 | 0.12 | 51.40% | 50.00% | 1.05 | 0.02 | 164.58 | 8.97 | 70.22 | 0.67 |
| RDH12 | 29 | 279 | 9 | 36 | 67 | 1.92 | 1.01 | 18.02% | 11.77% | 58.06% | 22.35 | 1.10 | 19.00% | 37.32% | 33.69% | 0.37 | 0.52 | 49.05% | 22.06% | 34.05% | 1.51 | 2.52 | 57.88% | 84.76% | 0.99 | 0.03 | 179.48 | 3.48 | 20.59 | 0.72 |
| RDH5 | 9 | 76 | 13 | 50 | 84 | 1.21 | 1.04 | 27.31% | 3.19E-05 | 22.37% | 2.90 | 0.39 | 16.27% | 17.58% | 23.68% | 0.14 | 0.38 | 44.14% | 8.08% | 19.74% | 0.25 | 0.58 | 41.17% | 40.00% | 1.01 | 0.02 | 193.80 | 5.48 | 69.47 | 0.70 |
| REEP6 | 3 | 43 | 39 | 44 | 62 | 2.45 | 1.02 | 18.13% | 0.00% | 44.19% | 1.78 | 1.02 | 15.53% | 15.83% | 9.30% | 0.01 | 0.12 | 61.28% | 50.00% | 90.70% | 2.38 | 5.53 | 57.55% | 97.92% | 1.18 | 0.02 | 184.26 | 11.54 | 415.76 | 0.75 |
| RGR | 1 | 10 | 85 | 85 | 85 | 1.95 | 1.00 | 31.35% | 0.00% | 100.00% | 111.10 | 3.50 | 33.96% | 17.84% | 0.00% | 0.00 | 0.00 | | | 0.00% | 0.00 | 0.00 | | | 1.14 | 0.02 | 218.95 | 6.17 | 66.37 | 0.73 |
| RHO | 107 | 968 | 9 | 50 | 95 | 2.17 | 1.03 | 15.53% | 1.53% | 61.88% | 9.74 | 1.72 | 13.64% | 34.76% | 18.49% | 0.11 | 0.24 | 52.95% | 55.98% | 70.56% | 2.25 | 2.99 | 53.62% | 82.80% | 1.22 | 0.04 | 175.72 | 6.49 | 86.13 | 0.69 |
| RLBP1 | 6 | 42 | 15 | 46.5 | 83 | 1.62 | 1.00 | 19.39% | 0.30% | 69.05% | 7.51 | 1.90 | 16.27% | 27.24% | 33.33% | 0.04 | 0.38 | 45.37% | 63.77% | 9.52% | 0.04 | 0.12 | 40.72% | 100.00% | 1.11 | 0.03 | 209.37 | 19.39 | 327.98 | 0.69 |
| RP1 | 115 | 951 | 18 | 59.5 | 93 | 2.19 | 1.01 | 13.69% | 5.55% | 62.99% | 9.32 | 1.76 | 13.07% | 40.06% | 19.14% | 0.18 | 0.27 | 55.90% | 66.70% | 64.98% | 2.03 | 2.98 | 54.56% | 74.81% | 1.23 | 0.05 | 174.37 | 6.93 | 139.44 | 0.70 |
| RP1L1 | 18 | 169 | 13 | 55 | 90 | 1.82 | 1.04 | 16.11% | 7.92% | 48.52% | 3.52 | 1.76 | 4.43% | 29.00% | 8.28% | 0.00 | 0.06 | 56.38% | 44.95% | 30.18% | 0.79 | 1.15 | 54.11% | 79.65% | 1.30 | 0.08 | 200.64 | 4.74 | 46.77 | 0.66 |
| RP2 | 28 | 332 | 11 | 31.5 | 81 | 2.48 | 1.05 | 9.87% | 13.25% | 47.59% | 3.10 | 1.19 | 10.55% | 48.28% | 6.93% | 0.02 | 0.08 | 54.74% | 84.40% | 12.95% | 0.26 | 0.38 | 58.97% | 67.79% | 1.19 | 0.04 | 181.19 | 6.83 | 109.26 | 0.69 |
| RP9 | 9 | 103 | 21 | 50 | 74 | 2.10 | 1.02 | 15.49% | 1.94% | 40.78% | 3.50 | 1.51 | 11.09% | 13.82% | 7.77% | 0.01 | 0.07 | 48.98% | 50.03% | 67.96% | 3.34 | 1.70 | 54.39% | 71.25% | 1.24 | 0.05 | 171.05 | 6.24 | 121.52 | 0.69 |
| RPE65 | 18 | 82 | 5 | 26 | 82 | 1.15 | 1.06 | 19.28% | 7.57% | 52.44% | 40.13 | 2.65 | 15.03% | 29.14% | 37.80% | 0.24 | 0.65 | 56.35% | 61.75% | 4.88% | 0.19 | 0.05 | 43.10% | 97.60% | 0.97 | 0.02 | 164.35 | 6.64 | 110.77 | 0.65 |
| RPGR | 161 | 1429 | 7 | 44 | 87 | 2.31 | 1.01 | 13.72% | 3.67% | 54.23% | 7.78 | 1.27 | 14.36% | 36.54% | 22.60% | 0.13 | 0.31 | 54.42% | 58.64% | 61.30% | 2.13 | 2.09 | 55.94% | 81.99% | 1.19 | 0.04 | 182.93 | 8.03 | 126.10 | 0.70 |
| RPGRIP1 | 7 | 93 | 17 | 27.5 | 59 | 2.73 | 1.00 | 13.28% | 0.00% | 4.30% | 0.00 | 0.04 | 7.57% | 25.00% | 15.05% | 0.02 | 0.15 | 51.47% | 0.00% | 70.97% | 1.08 | 3.42 | 43.87% | 85.01% | 1.16 | 0.03 | 206.41 | 9.28 | 284.07 | 0.71 |
| RS1 | 100 | 1186 | 7 | 33 | 78 | 1.93 | 1.00 | 11.86% | 3.50% | 26.81% | 2.41 | 0.58 | 10.68% | 36.67% | 24.79% | 0.10 | 0.37 | 44.28% | 73.17% | 16.95% | 0.35 | 0.59 | 53.00% | 66.36% | 1.34 | 0.09 | 207.50 | 5.45 | 62.27 | 0.72 |
| SAG | 4 | 47 | 15 | 47.5 | 58 | 2.99 | 1.04 | 13.28% | 0.00% | 31.91% | 1.19 | 0.53 | 20.98% | 63.99% | 0.00% | 0.00 | 0.00 | | | 14.89% | 0.42 | 0.47 | 54.36% | 89.84% | 1.20 | 0.04 | 228.97 | 9.02 | 87.09 | 0.69 |
| SDCCAG8 | 1 | 11 | 29 | 29 | 29 | 3.19 | 1.00 | 9.09% | 0.00% | 0.00% | 0.00 | 0.00 | | | 0.00% | 0.00 | 0.00 | | | 100.00% | 1.43 | 3.91 | 51.65% | 100.00% | 1.40 | 0.08 | 182.97 | 7.54 | 83.15 | 0.76 |
| SGSH | 1 | 2 | 34 | 34 | 34 | 3.23 | 1.00 | 6.49% | 0.00% | 100.00% | 2.46 | 2.50 | 6.39% | 0.15% | 0.00% | 0.00 | 0.00 | | | 100.00% | 1.38 | 3.00 | 40.38% | 100.00% | 1.32 | 0.05 | 188.58 | 6.61 | 32.76 | 0.73 |
| SLC24A1 | 1 | 4 | 28 | 28 | 28 | 2.67 | 1.00 | 13.92% | 0.00% | 0.00% | 0.00 | 0.00 | | | 75.00% | 0.92 | 3.75 | 67.49% | 0.00% | 75.00% | 1.77 | 1.75 | 50.43% | 0.00% | 1.46 | 0.11 | 179.34 | 4.31 | 51.46 | 0.73 |
| SLC24A5 | 1 | 5 | 18 | 21 | 28 | 1.75 | 1.00 | 13.56% | 0.00% | 0.00% | 0.00 | 0.00 | | | 0.00% | 0.00 | 0.00 | | | 0.00% | 0.00 | 0.00 | | | 1.36 | 0.10 | 242.22 | 10.66 | 162.99 | 0.76 |
| SLC25A46 | 1 | 4 | 65 | 65 | 65 | 2.15 | 1.00 | 17.71% | 0.00% | 0.00% | 0.00 | 0.00 | | | 0.00% | 0.00 | 0.00 | | | 0.00% | 0.00 | 0.00 | | | 1.44 | 0.11 | 195.15 | 6.97 | 144.31 | 0.71 |
| SNRNP200 | 12 | 122 | 29 | 48 | 79 | 2.26 | 1.00 | 19.33% | 0.20% | 73.77% | 18.05 | 2.05 | 15.78% | 29.08% | 30.33% | 0.21 | 0.52 | 51.82% | 69.42% | 74.59% | 2.13 | 3.57 | 49.38% | 96.55% | 1.13 | 0.03 | 166.44 | 6.64 | 88.10 | 0.69 |
| SPATA7 | 2 | 20 | 21 | 35 | 57 | 2.54 | 1.15 | 13.78% | 4.94% | 5.00% | 0.13 | 0.10 | 28.36% | 0.00% | 35.00% | 0.05 | 0.35 | 62.23% | 13.51% | 90.00% | 1.83 | 3.60 | 40.23% | 90.97% | 1.14 | 0.03 | 197.80 | 8.37 | 98.86 | 0.70 |
| SSBP1 | 3 | 35 | 29 | 62 | 63 | 1.38 | 1.00 | 16.74% | 18.76% | 77.14% | 0.62 | 2.49 | 8.56% | 49.16% | 20.00% | 0.01 | 0.26 | 76.85% | 0.00% | 40.00% | 1.41 | 1.86 | 54.33% | 69.36% | 1.20 | 0.04 | 165.92 | 5.26 | 92.93 | 0.73 |
| TIMP3 | 36 | 322 | 26 | 57 | 78 | 1.71 | 1.01 | 17.51% | 9.49% | 62.11% | 16.35 | 1.84 | 15.26% | 46.54% | 30.75% | 0.14 | 0.51 | 58.46% | 32.56% | 27.02% | 0.47 | 1.00 | 55.24% | 35.32% | 1.39 | 0.09 | 198.27 | 4.99 | 71.86 | 0.71 |
| TOPORS | 6 | 83 | 20 | 52.5 | 72 | 2.16 | 0.99 | 13.72% | 0.00% | 75.90% | 2.58 | 1.88 | 17.39% | 80.33% | 4.82% | 0.01 | 0.05 | 70.07% | 25.00% | 93.98% | 3.05 | 4.57 | 56.99% | 81.22% | 1.29 | 0.06 | 187.25 | 6.93 | 80.81 | 0.71 |
| TRNT1 | 1 | 10 | 41 | 41 | 41 | 2.62 | 1.00 | 15.62% | 0.00% | 10.00% | 0.00 | 0.00 | 13.96% | 0.00% | 0.00% | 0.00 | 0.00 | | | 100.00% | 5.69 | 3.60 | 70.63% | 94.80% | 1.15 | 0.02 | 184.94 | 10.53 | 269.64 | 0.68 |
| TRPM1 | 6 | 27 | 8 | 21 | 57 | 2.65 | 1.00 | 14.22% | 0.67% | 0.00% | 0.00 | 0.00 | | | 0.00% | 0.00 | 0.00 | | | 0.00% | 0.00 | 0.00 | | | 1.24 | 0.06 | 207.28 | 7.28 | 63.60 | 0.66 |
| TSPAN12 | 1 | 1 | 23 | 30.5 | 38 | 2.16 | 1.00 | 9.87% | 0.00% | 0.00% | 0.00 | 0.00 | | | 0.00% | 0.00 | 0.00 | | | 0.00% | 0.00 | 0.00 | | | 1.47 | 0.13 | 205.51 | 4.79 | 64.06 | 0.71 |
| TTLL5 | 8 | 56 | 37 | 47 | 70 | 2.34 | 1.00 | 14.86% | 1.59% | 85.71% | 3.36 | 1.68 | 17.98% | 95.01% | 30.36% | 0.03 | 0.27 | 37.89% | 100.00% | 100.00% | 3.07 | 5.32 | 50.65% | 88.34% | 1.32 | 0.06 | 213.57 | 8.00 | 121.73 | 0.76 |
| TULP1 | 16 | 165 | 7 | 33 | 69 | 2.57 | 1.05 | 13.77% | 2.78% | 26.06% | 0.53 | 0.41 | 15.25% | 63.18% | 21.82% | 0.07 | 0.40 | 52.38% | 27.71% | 78.18% | 2.56 | 2.96 | 47.46% | 75.10% | 1.15 | 0.04 | 199.61 | 12.24 | 251.64 | 0.70 |
| TYR | 12 | 91 | 5 | 34 | 76 | 1.63 | 1.00 | 12.26% | 12.36% | 3.30% | 0.01 | 0.14 | 17.37% | 16.77% | 9.89% | 0.01 | 0.20 | 63.87% | 26.92% | 0.00% | 0.00 | 0.00 | | | 1.37 | 0.09 | 217.99 | 5.12 | 69.35 | 0.72 |
| TYRP1 | 1 | 4 | 13 | 19.5 | 26 | 1.87 | 1.00 | 12.90% | 0.00% | 0.00% | 0.00 | 0.00 | | | 0.00% | 0.00 | 0.00 | | | 0.00% | 0.00 | 0.00 | | | 1.46 | 0.13 | 208.11 | 3.89 | 32.71 | 0.73 |
| USH1C | 14 | 99 | 22 | 41 | 81 | 2.05 | 1.01 | 19.20% | 2.54% | 66.67% | 7.01 | 1.63 | 11.29% | 34.47% | 29.29% | 0.60 | 0.56 | 52.39% | 41.64% | 46.46% | 1.83 | 1.59 | 46.45% | 62.73% | 1.01 | 0.02 | 167.35 | 14.95 | 516.55 | 0.67 |
| USH1G | 2 | 12 | 32 | 38.5 | 45 | 1.89 | 1.00 | 13.40% | 11.93% | 33.33% | 0.20 | 0.42 | 9.90% | 0.00% | 50.00% | 0.33 | 1.08 | 56.63% | 0.00% | 58.33% | 2.89 | 5.00 | 58.58% | 1.79% | 1.23 | 0.04 | 234.70 | 8.28 | 94.58 | 0.73 |
| USH2A | 306 | 2858 | 0 | 52 | 91 | 2.17 | 1.02 | 15.63% | 2.32% | 61.30% | 8.47 | 1.76 | 12.99% | 36.45% | 19.84% | 0.15 | 0.31 | 53.46% | 66.47% | 72.08% | 1.98 | 3.20 | 53.47% | 87.51% | 1.17 | 0.03 | 170.22 | 7.26 | 101.60 | 0.69 |
| USH2C | 5 | 59 | 47 | 56 | 61 | 1.69 | 0.98 | 16.12% | 3.42% | 88.14% | 10.99 | 2.12 | 11.67% | 20.83% | 32.20% | 0.24 | 0.47 | 58.05% | 90.79% | 79.66% | 2.09 | 2.98 | 52.11% | 88.24% | 1.07 | 0.02 | 156.45 | 6.12 | 82.23 | 0.70 |
| VHL | 1 | 2 | 27 | 27 | 27 | 2.78 | 1.00 | 20.14% | 0.00% | 0.00% | 0.00 | 0.00 | | | 0.00% | 0.00 | 0.00 | | | 0.00% | 0.00 | 0.00 | | | 1.40 | 0.10 | 223.00 | 6.09 | 43.62 | 0.71 |
| VPS13B | 3 | 37 | 34 | 35 | 65 | 2.10 | 1.00 | 25.51% | 0.00% | 48.65% | 1.62 | 0.57 | 19.57% | 85.24% | 18.92% | 0.04 | 0.22 | 43.57% | 4.28% | 70.27% | 1.74 | 1.30 | 49.23% | 94.07% | 0.85 | 0.01 | 172.54 | 9.77 | 192.76 | 0.76 |
| WDR19 | 2 | 27 | 18 | 31 | 44 | 2.57 | 1.00 | 12.46% | 0.00% | 22.22% | 2.93 | 0.41 | 14.56% | 92.55% | 11.11% | 0.00 | 0.11 | 30.38% | 92.59% | 96.30% | 1.55 | 2.37 | 57.59% | 20.26% | 1.37 | 0.07 | 162.48 | 3.79 | 22.94 | 0.75 |
| WFS1 | 4 | 21 | 18 | 32.5 | 79 | 1.87 | 1.00 | 24.03% | 1.16% | 0.00% | 0.00 | 0.00 | | | 19.05% | 0.01 | 1.10 | 62.50% | 21.98% | 0.00% | 0.00 | 0.00 | | | 1.40 | 0.10 | 208.53 | 4.40 | 52.44 | 0.74 |
